# Prostate cancer risk stratification via non-destructive 3D pathology with annotation-free gland segmentation and analysis

**DOI:** 10.1101/2021.08.30.21262847

**Authors:** W. Xie, N.P. Reder, C. Koyuncu, P. Leo, S. Hawley, H. Huang, C. Mao, N. Postupna, S. Kang, R. Serafin, G. Gao, Q. Han, K.W. Bishop, L.A. Barner, P. Fu, J.L. Wright, C.D. Keene, J.C. Vaughan, A. Janowczyk, A.K. Glaser, A. Madabhushi, L.D. True, J.T.C. Liu

## Abstract

Prostate cancer treatment planning is largely dependent upon examination of core-needle biopsies. In current clinical practice, the microscopic architecture of the prostate glands is what forms the basis for prognostic grading by pathologists. Interpretation of these convoluted 3D glandular structures via visual inspection of a limited number of 2D histology sections is often unreliable, which contributes to the under- and over-treatment of patients. To improve risk assessment and treatment decisions, we have developed a workflow for non-destructive 3D pathology and computational analysis of whole prostate biopsies labeled with a rapid and inexpensive fluorescent analog of standard H&E staining. Our analysis is based on interpretable glandular features, and is facilitated by the development of image-translation-assisted segmentation in 3D (ITAS3D). ITAS3D is a generalizable deep-learning-based strategy that enables tissue microstructures to be volumetrically segmented in an annotation-free and objective (biomarker-based) manner without requiring real immunolabeling. To provide evidence of the translational value of a computational 3D pathology approach, we analyzed *ex vivo* biopsies (*n* = 300) extracted from archived radical-prostatectomy specimens (*N* = 50), and found that 3D glandular features are superior to corresponding 2D features for risk stratification of low-to intermediate-risk PCa patients based on their clinical biochemical recurrence (BCR) outcomes.

**Significance:** We present an end-to-end pipeline for computational 3D pathology of whole prostate biopsies, showing that non-destructive pathology has the potential to enable superior prognostic stratification for guiding critical oncology decisions.

## Introduction

Prostate cancer (PCa) is the most common cancer in men and the second leading cause of death for men in the United States ^1^. Currently, PCa management is largely dependent upon examination of prostate biopsies via 2D histopathology ^2^, in which a set of core-needle biopsies is formalin-fixed and paraffin-embedded (FFPE) to allow thin sections to be cut, mounted on glass slides, and stained for microscopic analysis. To quantify the aggressiveness of the cancer, the Gleason grading system is used, which relies entirely upon visual interpretation of prostate gland morphology as seen on 2D histology slides. Unfortunately, Gleason grading of PCa is associated with high levels of interobserver variability ^3,4^ and is only moderately correlated with outcomes, especially for patients with intermediate-grade PCa ^5^. This contributes to the undertreatment of patients with aggressive cancer (e.g. with active surveillance) ^6^, leading to preventable metastasis and death ^7^, and the overtreatment of patients with indolent cancer (e.g. with surgery or radiation therapy) ^8^, which can lead to serious side effects, such as incontinence and impotence ^9^.

With conventional histopathology, only a small fraction of a clinical specimen (e.g., a biopsy) is typically sampled, and key prognostic microstructures are viewed as 2D cross-sections rather than in their native 3D configurations ^10^. Here, the term “sampled” refers to the volume of an excised specimen that is converted into slide-mounted histology sections and viewed by a pathologist. For example, for a typical 1-mm diameter needle biopsy, only a few 4-μm-thick sections are visualized, representing ∼1% of the biopsy. Attempting to interpret complex and heterogeneous 3D tissue structures using thin 2D tissue sections results in ambiguities and artifacts. For example, well-formed prostate carcinoma glands (Gleason pattern 3) can appear to be poorly formed (Gleason pattern 4) if those glands are tangentially sectioned on a 2D slide, which can have major implications on disease management (e.g., active surveillance vs. surgery / radiation) ^11-13^.

Motivated by recent technological advances in optical clearing to render tissue specimens transparent to light (i.e. iDISCO ^14^, CUBIC ^15^ etc.) in conjunction with high-throughput 3D light-sheet microscopy, a number of groups have been exploring the value of non-destructive 3D pathology of clinical specimens for diagnostic pathology ^13,16-20^. Compared to conventional slide-based histology, non-destructive 3D pathology can achieve vastly greater sampling of large specimens along with volumetric visualization and quantification of diagnostically significant microstructures, all while maintaining intact specimens for downstream molecular assays ^10^. However, since the associated information content of a 3D pathology dataset of a biopsy is >100× larger than a 2D whole-slide image representation (in terms of total number of pixels), computational tools are necessary to analyze these large datasets efficiently and reproducibly for diagnostic and prognostic determinations.

We hypothesized that 3D vs. 2D pathology datasets could allow for improved characterization of the convoluted glandular structures that pathologists currently rely on for PCa risk stratification. A multi-stage computational pipeline was desired for classifying patient outcomes based on interpretable “hand-crafted” features ^21-23^ (i.e., glandular features) rather than an end-to-end deep-learning (DL) strategy for risk classification based directly on the imaging data ^24-26^. This was motivated by: (1) the attractiveness of an intuitive feature-based approach as an initial strategy to facilitate hypothesis-testing and clinical adoption of an emerging modality in which datasets are currently limited and poorly understood ^10,27^, and (2) the observation that when case numbers are limited, a hand-crafted feature-based approach can be more reliable than an end-to-end DL classifier ^28,29^.

Multi-stage feature-based classification approaches rely on the accurate segmentation of morphological structures such as nuclei ^30,31^, collagen fibers ^32,33^, vessels ^18,34^, or in our case, prostate glands ^21,35^. This is typically achieved in one of two ways: (1) Direct DL-based segmentation methods ^36-39^ that require manually annotated training datasets, which are especially tedious and difficult to obtain in 3D (**Fig. 1a**) ^40^; or (2) traditional computer vision (CV) approaches based on intensity and morphology, provided that tissue structures of interest can be stained/labeled with high specificity (**Fig. 1b**) ^41,42^. While immunolabeling can confer a high degree of specificity for traditional CV-based segmentation, it is not an attractive strategy for clinical 3D pathology assays due to the high cost of antibodies to stain large tissue volumes, and the slow diffusion times of antibodies in thick tissues (up to several weeks) ^14,43^.

**Fig. 1.**
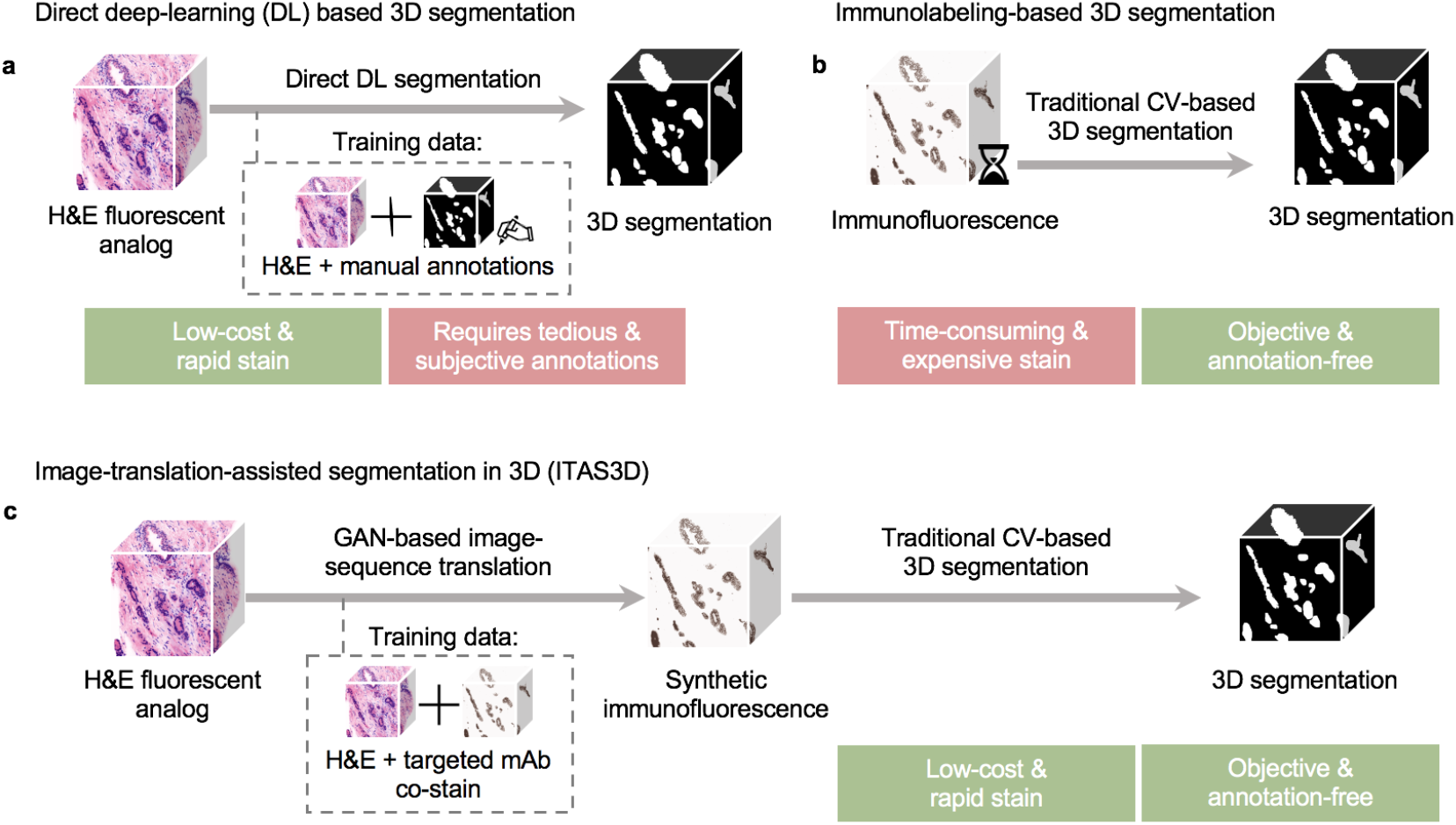
General methods for 3D gland segmentation. **a**, A single-step deep learning (DL) segmentation model can be trained with imaging datasets of tissues labeled with a fluorescent analog of H&E paired with manually annotated ground-truth segmentation masks. While H&E-analog staining is low-cost and rapid, manual annotations are labor-intensive (especially in 3D) and based on subjective human judgements. **b**, By immunolabeling a tissue microstructure with high specificity, 3D segmentations can be achieved with traditional computer-vision (CV) methods without the need for manual annotations. While this is an objective segmentation method based on a chemical biomarker, immunolabeling large intact specimens is expensive and time-consuming due to the slow diffusion of antibodies in thick tissues. **c**, With image-translation-assisted segmentation in 3D (ITAS3D), H&E-analog datasets are computationally transformed in appearance to mimic immunofluorescence datasets, which enables the synthetically labeled tissue structures to be segmented with traditional CV methods. The image-sequence translation model is trained with a generative adversarial network (GAN) based on paired H&E-analog and immunofluorescence datasets. ITAS3D is rapid and low-cost (in terms of staining) as well as annotation-free and objective (i.e. biomarker-based).

To address these challenges, we developed a generalizable annotation-free 3D segmentation method, hereafter referred to as “image-translation-assisted segmentation in 3D (ITAS3D)”. In our specific implementation of ITAS3D (**Fig. 1c**), 3D H&E-analog images of prostate tissues are synthetically converted in appearance to mimic 3D immunofluorescence (IF) images of Cytokeratin 8 (CK8) – a low-molecular-weight keratin expressed by the luminal epithelial cells of all prostate glands – thereby facilitating the objective (biomarker-based) segmentation of the glandular epithelium and lumen spaces using traditional CV tools. The deep-learning image-translation algorithm is trained with a generative adversarial network (GAN), which has been previously used for 2D virtual-staining applications ^44-46^. However, unlike those prior 2D image-translation efforts, we developed a “2.5D” virtual-staining approach based on a specialized GAN that was originally designed to achieve video translation with high spatial continuity between frames, but which has been adapted within our ITAS3D framework to ensure high spatial continuity as a function of depth (see Results) ^47^.

As a clinical study to investigate the value of our computational 3D pathology workflow, 300 *ex vivo* biopsies were extracted from archived radical prostatectomy (RP) specimens obtained from 50 patients who underwent surgery over a decade ago. We stained the biopsies with an inexpensive small-molecule (i.e., rapidly diffusing) fluorescent analog of H&E, optically cleared the biopsies with a simple dehydration and solvent-immersion protocol to render them transparent to light, and then used an open-top light-sheet (OTLS) microscopy platform to obtain whole-biopsy 3D pathology datasets. The prostate glandular network was then segmented using ITAS3D, from which 3D glandular features (i.e., histomorphometric parameters) and corresponding 2D features were extracted. These 3D and 2D features were evaluated for their ability to stratify patients based on clinical biochemical recurrence (BCR) outcomes, which serve as a proxy endpoint for aggressive vs. indolent PCa.

## Materials and Methods

Key methodological information is provided below for the clinical-validation study. The Supplementary Methods contain technical details on the training and validation of ITAS3D, as well as computational details for the clinical study, such as a description of gland features.

### Collection and processing of archived tissue to obtain simulated biopsies for the clinical study

Archived FFPE prostatectomy specimens were collected from 50 PCa patients (see **Supplementary Table 3** for clinical data) of which 46 cases were initially graded during post-RP histopathology as having Gleason scores of 3+3, 3+4 or 4+3 (Grade Group 1 – 3). All patients were followed up for at least 5 years post-RP as part of a prior study (Canary TMA) ^57^. For deparaffinization, the identified FFPE blocks were first heated at 75°C for 1 hour until the outer paraffin wax was melted, and then treated with 65°C xylene for 48 hours. Next, one simulated core-needle biopsy (∼1-mm in width) was cut from each of the 6 deparaffinized blocks (per patient case), resulting in a total of n = 300 biopsy cores. All simulated biopsies were then fluorescently labeled with the T&E version of our H&E-analog staining protocol.

### The T&E staining protocol (H&E analog) for the clinical study

Biopsies were first washed in 100% ethanol twice for 1 h each to remove any excess xylene, then treated in 70% ethanol for 1 h to partially re-hydrate the biopsies. Each biopsy was then placed in an individual 0.5ml Eppendorf tube (Cat: 14-282-300, Fisher Scientific), stained for 48 hours in 70% ethanol at pH 4 with a 1:200 dilution of Eosin-Y (Cat: 3801615, Leica Biosystems) and a 1:500 dilution of To-PRO™-3 Iodide (Cat: T3605, Thermo-Fisher) at room temperature with gentle agitation. The biopsies were then dehydrated twice in 100% ethanol for 2 hours. Finally, the biopsies were optically cleared (n = 1.56) by placing them in ethyl cinnamate (Cat: 112372, Sigma-Aldrich) for 8 hours before imaging them with open-top light-sheet (OTLS) microscopy.

### Open-top light-sheet (OTLS) microscopy and pre-processing

We utilized a previously developed OTLS microscope ^19^ to image tissues slices (for training data) and simulated biopsies (for the clinical study). For this study, ethyl cinnamate (n = 1.56) was used as the immersion medium, and a custom-machined HIVEX plate (n=1.55) was used as a multi-biopsy sample holder (12 biopsies per holder). Multi-channel illumination was provided by a four-channel digitally controlled laser package (Skyra, Cobolt Lasers). Tissues were imaged at near-Nyquist sampling of 0 ∼0.44 μm/pixel. The volumetric imaging time was approximately 0.5 min per mm^3^ of tissue for each wavelength channel. This allowed each biopsy (∼1 × 1 × 20 mm), stained with two fluorophores (T&E), to be imaged in ∼20 min.

### Statistical analysis of the correlation between glandular features and BCR outcomes

Patient-level glandular features were obtained by averaging the biopsy-level features from all cancer-containing biopsies from a single patient. Patients who experienced BCR within 5 years post-RP are denoted as the “BCR” group, and all other patients are denoted as “non-BCR”. BCR was defined here as a rise in serum levels of prostate specific antigen (PSA) to 0.2 ng/ml after 8 weeks post-RP ^57^. The box plots indicate median values along with interquartile ranges (25% - 75% of the distribution). The whiskers extend to the furthest data points excluding outliers defined as points beyond 1.5× the interquartile range. The *p* values for the BCR group vs. non-BCR group are calculated using the two-sided Mann–Whitney U-test ^82^. To assess the ability of different 3D and 2D glandular features to distinguish between BCR vs. non-BCR groups, we applied ROC curve analysis, from which an area-under-the-curve (AUC) value could be extracted. The *t*-SNE^83^ analyses were performed with 1000 iterations at a learning rate of 100.

To develop multiparameter classifiers to stratify patients based on 5-year BCR outcomes, a least absolute shrinkage and selection operator (LASSO) logistic regression model was developed ^22^ using the binary 5-year BCR category as the outcome endpoint. LASSO is a linear regression model that includes a L1 regularization term to avoid overfitting and to identify a subset of features that are most predictive. Here, the optimal LASSO tuning parameter, λ, was determined with 3-fold cross validation (CV), where the dataset was randomly partitioned into three equal-sized groups: two groups to train the model with a specific λ, and one group to test the performance of the model. Along the LASSO regularization path, the λ with the highest R^2^ (coefficient of determination) was defined as the optimal λ. Due to the lack of an external validation set, a nested CV schema was used to evaluate the performance of the multivariable models without any bias and data leakage between parameter estimation and validation steps^84^. The aforementioned CV used for hyperparameter tuning was performed in each iteration of the outer CV. LASSO regression was applied on the training set of the outer CV once an optimal λ was identified in the inner CV. AUC values were then calculated from the testing group of the outer CV (**Supplementary Fig. 8**). This nested CV was performed 200 times in order to determine an AUC (average and standard deviation). The exact same pipeline was used to develop multiparameter classifiers based on 3D and 2D features.

Kaplan Meier (KM) analysis was carried out to compare BCR-free survival rates for high-risk vs. low-risk groups of patients. This analysis utilized a subset of 34 cases for which time-to-recurrence data is available (see **Supplementary Table 3**). The performance of the models, either based on 2D or 3D features (non-skeleton features only), was quantified with *p* values (by log-rank test), hazard ratios (HR) and concordance index (C-index) metrics. For the multiparameter classification model used for KM analysis, the outer CV (3-fold) in our nested CV schema was replaced by a leave-one-out approach, where one case was left out of each iteration (50 total iterations) to calculate the probability of 5-year BCR for that patient ^85^. The samples were categorized as low- or high-risk by setting a posterior class probability threshold of 0.5. MATLAB was used for the KM analysis and all other statistical analysis was performed in Python with the “Scipy” and “Scikit-learn” packages.

## Results

### Annotation-free 3D gland segmentation

To segment the 3D glandular network within prostate biopsies, we first trained a GAN-based image-sequence translation model (see Supplementary Methods) to convert 3D H&E-analog images into synthetic CK8 IF images, which can be false colored to resemble chromogenic immunohistochemistry (IHC) (**Fig. 2**). As mentioned, the CK8 biomarker is expressed by the luminal epithelial cells of all prostate glands. The image-translation model is trained in a supervised manner with images from prostate tissues that are fluorescently tri-labeled with our H&E analog and a CK8-targeted monoclonal antibody (mAb).

**Fig. 2.**
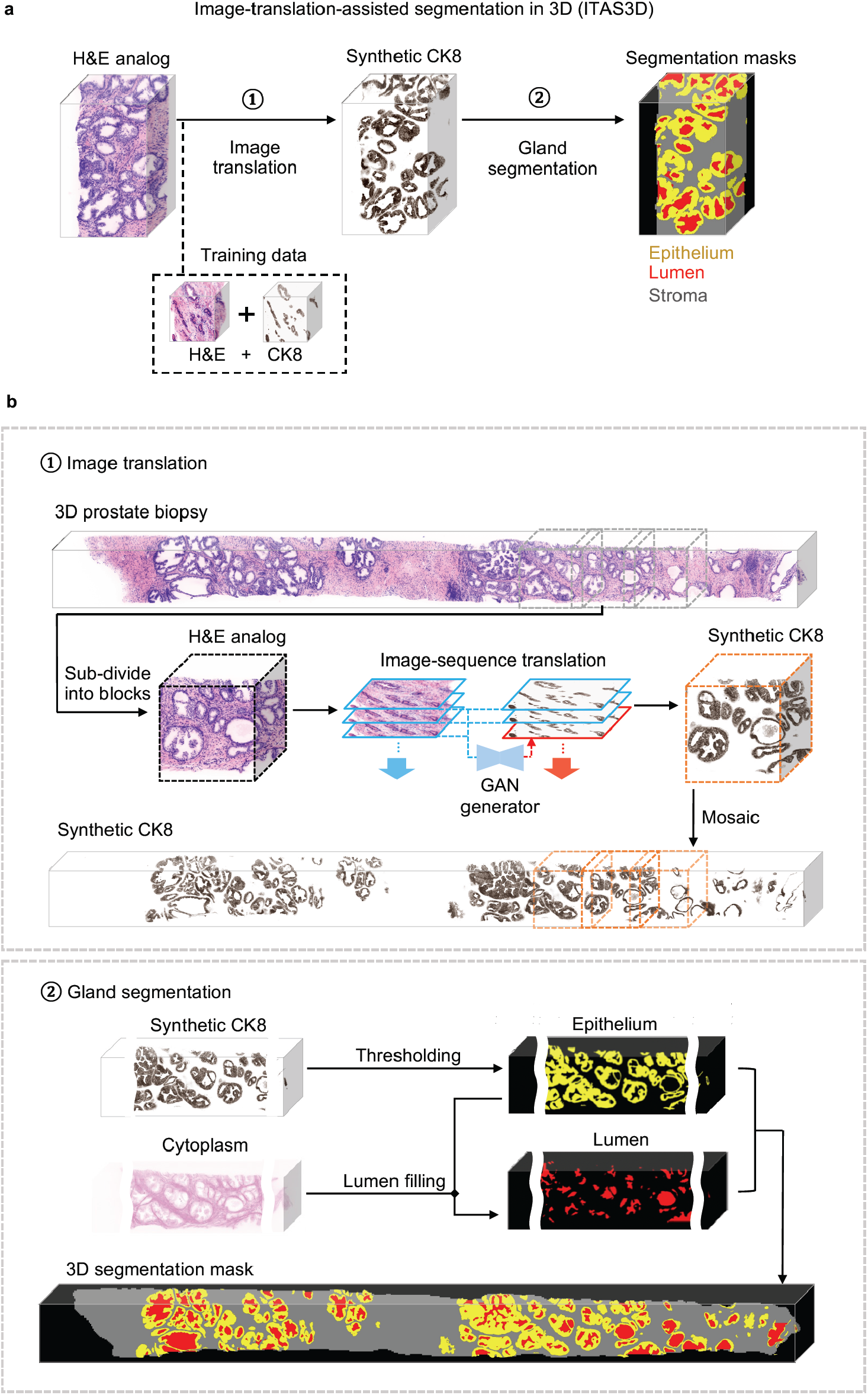
Image-translation-assisted segmentation in 3D (ITAS3D): a two-step pipeline for annotation-free 3D segmentation of prostate glands. **a**, In step 1, a 3D microscopy dataset of a specimen, stained with a rapid and inexpensive fluorescent analog of H&E, is converted into a synthetic CK8 immunofluorescence (IF) dataset by using an image-sequence translation model that is trained with paired H&E-analog and real-CK8 IF datasets (tri-labeled tissues). The CK8 biomarker, which is utilized in standard-of-care genitourinary pathology practice, is ubiquitously expressed by the luminal epithelial cells of all prostate glands. In step 2, traditional computer-vision algorithms are applied to the synthetic-CK8 datasets for semantic segmentation of the gland epithelium, lumen, and surrounding stromal regions. **b**, In step 1, a 3D prostate biopsy is sub-divided into overlapping blocks that are each regarded as depth-wise sequences of 2D images. A GAN-trained generator performs image translation sequentially on each 2D level of an image block. The image translation at each level is based on the H&E-analog input at that level while leveraging the H&E-analog and CK8 images from two previous levels to enforce spatial continuity between levels (i.e., a “2.5D” translation method). The synthetic-CK8 image-block outputs are then mosaicked to generate a 3D CK8 dataset of the whole biopsy to assist with gland segmentation. In step 2, the epithelial cell layer (epithelium) is segmented from the synthetic-CK8 dataset with a thresholding-based algorithm. Gland lumen spaces are segmented by filling in the regions enclosed by the epithelia with refinements based on the cytoplasm channel (eosin fluorescence). See Supplementary Methods for details.

As shown in step 1 of **Fig. 2**, for whole-biopsy H&E-to-CK8 conversion, we first sub-divide the 3D biopsy (∼ 1 mm × 0.7 mm × 20 mm) datasets into ∼1 mm × 0.7 mm × 1 mm (∼ 1024 × 700 × 1024 pixel) blocks. Each 3D image block is treated as a 2D image sequence as a function of depth. At each depth level, a synthetic CK8 image is inferred from the H&E-analog image at that level while simultaneously utilizing the images (H&E analog and CK8) from two previous levels to enforce spatial continuity as a function of depth. This “2.5D” image translation method is based on a previously reported “*vid2vid*” method for video translation (time sequences rather than depth sequences) ^47^ (see **Supplementary Fig. 1** and Supplementary Methods). However, our modified model omits the “coarse-to-fine” training strategy implemented in the original *vid2vid* method because this enables training times to be minimized with negligible performance loss (see **Supplementary Note 1, Supplementary Fig. 2** and **Supplementary Video 1**). Once the synthetic CK8 image blocks are generated, they are mosaicked to generate a whole-biopsy CK8 IHC dataset for gland segmentation. In step 2 of **Fig. 2**, the synthetic CK8 dataset is used to segment the luminal epithelial cell layer (“Epithelium” in **Fig. 2**) via a thresholding algorithm. The gland-lumen space, which is enclosed by the epithelium layer, can then be segmented by utilizing both the epithelium segmentation mask and the cytoplasmic channel (eosin-analog images). Algorithmic details are provided in the Supplementary Methods.

### Evaluation of image translation and segmentation

Example 3D prostate gland-segmentation results are shown for benign and cancerous regions in **Fig. 3a**. While the glands can be delineated on the H&E-analog images by a trained observer, automated computational segmentation of the glands remains challenging ^49,50^. Here we demonstrate that 3D image translation based on H&E-analog inputs results in synthetic-CK8 outputs in which the luminal epithelial cells are labeled with high contrast and spatial precision. We further show that these synthetic-CK8 datasets allow for relatively straightforward segmentation of the gland epithelium, lumen, and surrounding stromal tissue compartments (**Fig. 3a**). Glands from various PCa subtypes are successfully segmented as shown in **Fig. 3a**, including two glandular patterns that are typically associated with low and intermediate risk, respectively: small discrete well-formed glands (Gleason 3) and cribriform glands consisting of epithelial cells interrupted by multiple punched-out lumina (Gleason 4). **Supplementary Video 2** shows depth sequences of an H&E-analog dataset, a synthetic-CK8 dataset, and a segmentation mask of the two volumetric regions shown in **Fig. 3a**. A whole-biopsy 3D segmentation is also depicted in **Supplementary Video 3**.

**Fig. 3.**
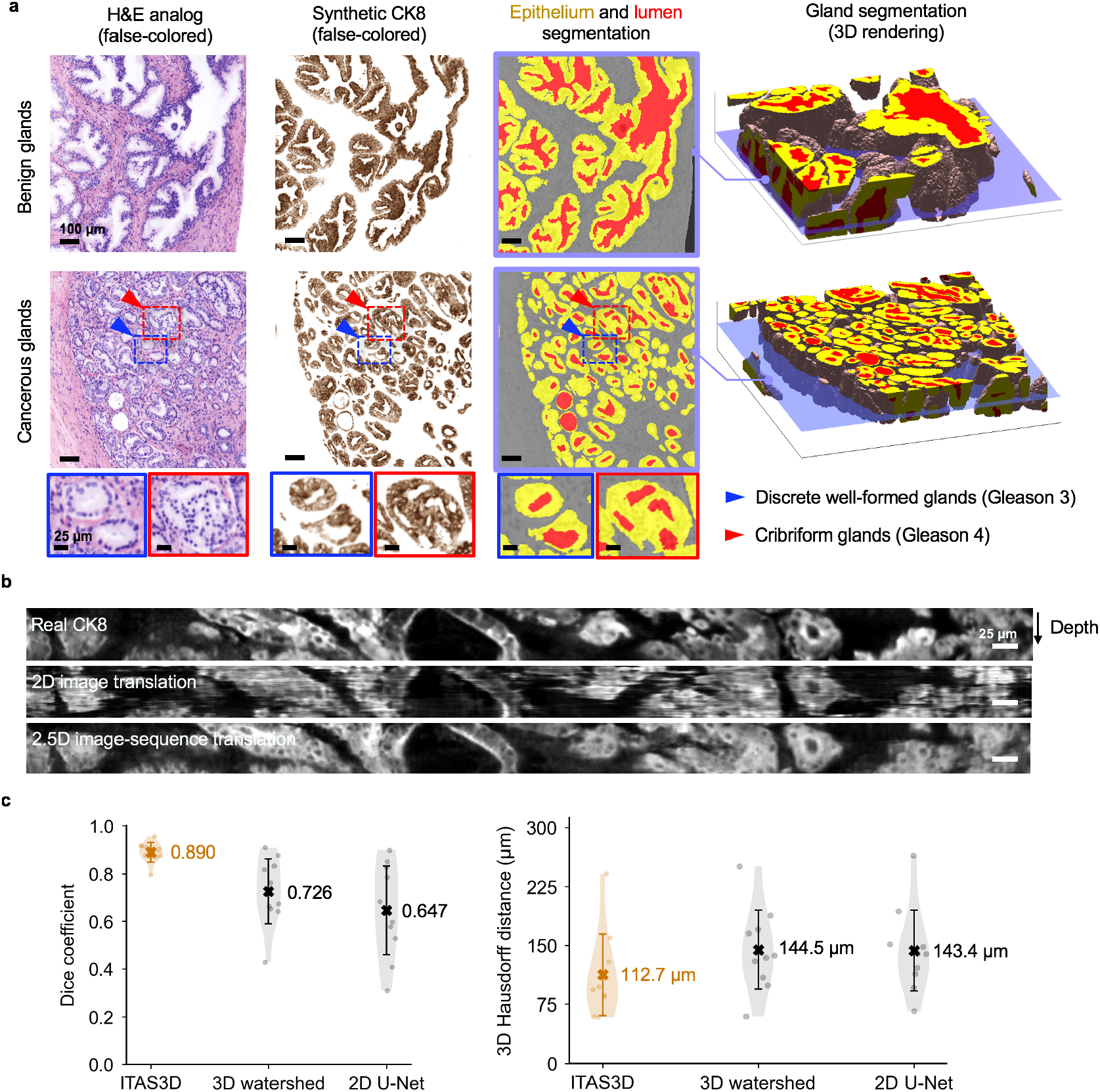
Segmentation results with ITAS3D. **a**, 2D cross-sections are shown (from left to right) of false-colored H&E-analog images, synthetic-CK8 IHC images generated by image-sequence translation, and gland-segmentation masks based on the synthetic-CK8 images (yellow for epithelium, red for lumen, and gray for stroma). The example images are from large 3D datasets containing benign glands (first row) and cancerous glands (second row). Zoom-in views show small discrete well-formed glands (Gleason pattern 3, blue box) and cribriform glands (Gleason pattern 4, red box) in the cancerous region. 3D renderings of gland segmentations, for a benign and cancerous region, are shown on the far right. Scale bar: 100 μm. **b**, Side views of the image sequences (with the depth direction oriented down) of real- and synthetic-CK8 immunofluorescence (IF) images. The 2.5D image-translation results exhibit substantially improved depth-wise continuity compared to the 2D image-translation results. Scale bar: 25 μm. **c**, For quantitative benchmarking, Dice coefficients (larger is better) and 3D Hausdorff distances (smaller is better) are plotted for ITAS3D-based gland segmentations along with two benchmark methods (3D watershed and 2D U-net), as calculated from 10 randomly selected test regions. Violin plots are shown with mean values denoted by a center cross and standard deviations denoted by error bars. For the 3D Hausdorff distance, the vertical axis denotes physical distance (in microns) within the tissue.

To demonstrate improved depth-wise continuity with our 2.5D image-translation strategy versus a similar 2D image-translation method (based on the “*pix2pix*” GAN), vertical cross-sectional views of a synthetic-CK8 dataset are shown in **Fig. 3b**. While obvious distortions and discontinuities are seen as a function of depth with 2D image translation, the results of our 2.5D image-sequence translation exhibit optimal continuity with depth. To further illustrate this improved continuity with depth, **Supplementary Video 4** shows a depth sequence of *en face* images (z stack). Abrupt morphological discontinuities between levels are again obvious with 2D translation but absent with the 2.5D translation approach. To quantify the performance of our image-translation method, a 3D structural similarity (SSIM) metric was calculated in which real CK8 IF datasets were used as ground truth. For images generated with 2.5D vs. 2D image translation, the 3D SSIM (averaged over 58 test volumes that were 0.2-mm^3^ each) was 0.41 vs. 0.37, reflecting a 12% improvement at a *p* value of 7.8 × 10^−6^ (two-sided paired t-test). This enhanced image-translation performance facilitates accurate 3D gland segmentations in subsequent steps of our computational pipeline.

To assess segmentation performance, ground-truth gland-segmentation datasets were first generated under the guidance of board-certified genitourinary pathologists (L.D.T. and N.P.R.). A total of 10 tissue volumes from different patients (512 × 512 × 100 pixels each, representing 0.2-mm^3^ of tissue) were manually annotated. We then compared the accuracy of ITAS3D with that of two common methods: 3D watershed ^51^ (as a 3D non-DL benchmark) and 2D U-Net ^52^ (as a 2D DL benchmark). ITAS3D outperforms the two benchmark methods in terms of Dice coefficient ^53^ and 3D Hausdorff distance ^54^ (**Fig. 3c)**. As a visual comparison between ITAS3D and the two benchmark methods, **Supplementary Video 5** displays image-stack sequences from three orthogonal perspectives of a representative segmented dataset, where the higher segmentation accuracy of ITAS3D can be appreciated. Note that a 3D DL-based benchmark method is not provided since there are currently insufficient 3D-annotated prostate gland datasets to train an end-to-end 3D DL segmentation model ^39,55,56^; again, this is one of the main motivations for developing the annotation-free ITAS3D method.

### Clinical validation study: glandular feature extraction and correlation with BCR outcomes

Due to the slow rate of progression for most PCa cases, an initial clinical study to assess the prognostic value of 3D vs. 2D glandular features was performed with archived prostatectomy specimens. Our study consisted of N = 50 PCa patients who were followed up for a minimum of 5 years post-RP as part of the Canary TMA case-cohort study ^57^. The Canary TMA study was based on a well-curated cohort of PCa patients in which the primary study endpoints were 5-year BCR outcomes and time to recurrence, which are also used as endpoints for our validation study. In the original Canary TMA study, approximately half of the patients experienced BCR within 5 years of RP, making it an ideal cohort for our study. We randomly selected a subset of 25 cases that had BCR within 5 years of RP (“BCR” group), and 25 cases that did not have BCR within 5 years of RP (“non-BCR” group).

FFPE tissue blocks were identified from each case corresponding to the 6 regions of the prostate targeted by urologists when performing standard sextant and 12-core (2 cores per sextant region) biopsy procedures (**Fig. 4a)**. Next, a simulated core-needle biopsy was extracted from each of the 6 FFPE tissue blocks for each patient (n = 300 total biopsy cores). The biopsies were deparaffinized, labeled with a fluorescent analog of H&E, optically cleared, and imaged nondestructively with a recently developed OTLS microscope ^19^ (see Methods). Review of the 3D pathology datasets by pathologists (L.D.T. and N.P.R.) revealed that 118 out of the 300 biopsy cores contained cancer (1 – 5 biopsies per case). The ITAS3D pipeline was applied to all cancer-containing biopsies. We then calculated histomorphometric features from the 3D gland segmentations, and from individual 2D levels from the center region of the biopsy cores (to mimic standard 2D pathology practices), which were then analyzed in terms of their association with BCR outcomes.

**Fig. 4.**
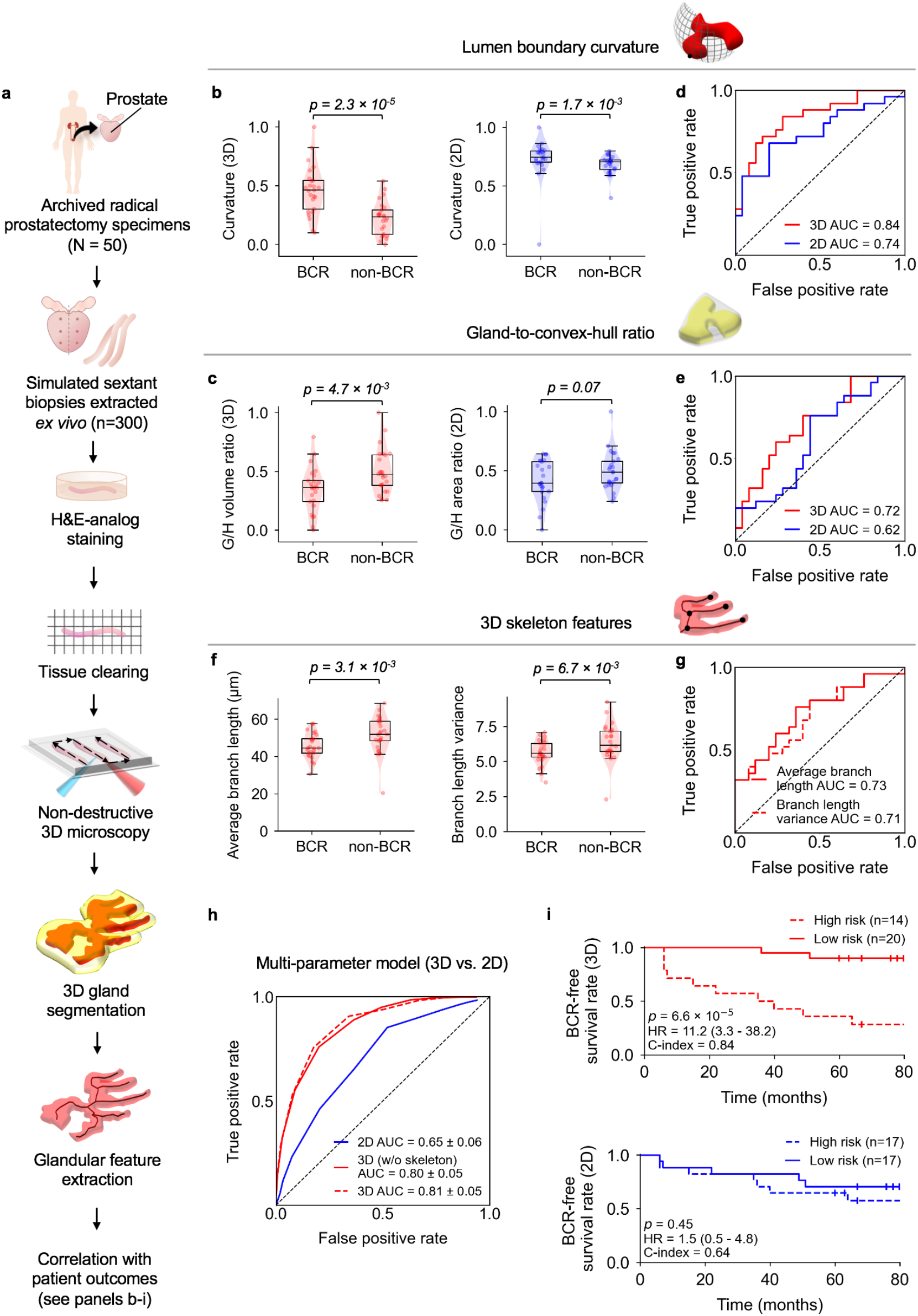
Clinical study comparing the performance of 3D vs. 2D glandular features for risk stratification. **a**, Archived (FFPE) radical prostatectomy (RP) specimens were obtained from a well-curated cohort of 50 patients treated over a decade ago, from which 300 simulated (*ex vivo*) needle biopsies were extracted (6 biopsies per case, per sextant-biopsy protocol). The biopsies were labeled with a fluorescent analog of H&E staining, optically cleared to render the tissues transparent to light, and then comprehensively imaged in 3D with open-top light-sheet (OTLS) microscopy. Prostate glands were computationally segmented from the resultant 3D biopsy images using the ITAS3D pipeline, and glandular features (shown in panels **b-g**) were extracted. **b**,**c**, Violin and box plots are shown for two examples of 3D glandular features, along with analogous 2D features, for cases in which biochemical recurrence (BCR) was observed within 5 years of RP (“BCR”) and for cases with no BCR within 5 years of RP (“non-BCR”). For both sets of example features, “lumen boundary curvature” in panel **b** and “gland-to-convex hull ratio” (G/H) in panel **c**, the 3D version of the feature shows improved stratification between BCR and non-BCR groups. **d**,**e**, Receiver-operating-characteristic (ROC) curves also show improved risk stratification with the 3D features vs. corresponding 2D features, with considerably higher area-under-the-curve (AUC) values. **f**, Violin and box plots are shown of representative gland-skeleton features (average branch length and branch length variance), which can only be accurately derived from the 3D pathology datasets, showing significant stratification between BCR and non-BCR groups. **g**, ROC curves are shown, along with AUC values, for average branch length and branch length variance. **h**, ROC curves are shown of various multiparameter models, including those trained with 2D glandular features, 3D glandular features excluding skeleton features, and 3D glandular features including skeleton features. **i**, Kaplan-Meier curves are shown for BCR-free survival, showing that a multiparameter model based on 3D glandular features is better able to stratify patients into low-risk and high-risk groups with significantly different recurrence trajectories (*p* = 6.6 × 10^−5^, HR = 11.2, C-index = 0.84).

We compared multiple 3D and 2D glandular histomorphometric features (see **Supplementary Table 1** for a detailed list). For example, the curvature of the boundary between the lumen and epithelium is a feature that increases as glands become smaller or more irregular, as is often seen with aggressive PCa ^21^. This can be quantified in the form of the average surface curvature of the object in 3D, or the curvature of the object’s cross-sectional circumference in 2D (**Fig. 4b**). As another example (**Fig. 4c**), the gland-to-convex-hull ratio (G/H) is defined as the volume ratio (in 3D) or the area ratio (in 2D) of the gland mask (epithelium + lumen) divided by the convex hull that circumscribes the gland. This G/H feature is inversely related to the irregularity or “waviness” of the periphery of the gland (at the scale of the gland itself rather than fine surface texture), which is generally expected to increase with aggressive PCa ^21^. For various 3D and 2D features (**Fig. 4d - 4e** and **Supplementary Table 1**), receiver operating characteristic (ROC) curves were generated to quantify the ability of the features to stratify patients based on 5-year BCR outcomes. When comparing analogous 3D and 2D glandular features, the 3D features largely exhibit an improved correlation with 5-year BCR outcomes in comparison to their 2D counterparts. This is exemplified by the significant *p* values for the 3D features showcased in **Fig. 4b – 4c** (between BCR and non-BCR groups) and higher area-under-the-ROC-curve (AUC) values (**Figs. 4d – 4e**).

We also extracted the 3D skeleton of the lumen network and quantified its branching parameters (skeleton-derived features). Example skeleton networks for benign and cancerous glands are shown in **Supplementary Video 6**. Due to the complex 3D branching-tree architecture of the gland-lumen network, there are no straightforward 2D analogs for these skeleton-derived features. In **Fig. 4d**, we show two examples of skeleton-derived features: the average branch length and the variance of the branch lengths. Both features are correlated with BCR outcomes based on *p* values and AUC values (**Fig. 4f – 4g**). Our analysis reveals that aggressive cancers (BCR cases) have shorter branch lengths and a smaller variance in branch lengths, which agrees with prior observations from 2D histology that glandular structures in higher-grade PCa are smaller and more abundant (i.e., less differentiated and varied in size). A histogram of branch lengths (**Supplementary Fig. 3)** demonstrates that the vast majority of branches are < 200-μm long, which suggests that the diameter of standard prostate biopsies (∼1-mm) is sufficient for whole-biopsy 3D pathology to quantify PCa branch lengths with reasonable accuracy.

To explore the prognostic value of combining multiple glandular features, we used linear regression models for feature selection and classification based on 3D vs. 2D features (see Methods). Brief descriptions and AUC values for the 3D and 2D glandular features involved in training the multi-parameter models are shown in **Supplementary Table 1**. The ROC curve of a model that combined 12 non-skeleton 3D features (“3D non-skeleton model”) yielded an AUC value of 0.80 ± 0.05 (average ± standard deviation; **Fig. 4h**), which is considerably higher than the AUC value (0.65 ± 0.06) of the model trained with 12 analogous 2D features (“2D model”). By adding 5 skeleton-derived features to the 12 non-skeleton 3D features, a re-trained 3D multiparameter model (“3D model”) yielded a slightly higher AUC value of 0.81 ± 0.05. The distribution of the 50 cases, based on their glandular features, can be visualized using *t*-distributed stochastic neighbor embedding (*t*-SNE), where a clearer separation between BCR and non-BCR cases is evident based on 3D vs. 2D glandular features (**Supplementary Fig. 4**). Multiparameter classification models based on 3D features alone (non-skeleton) or 2D features alone were used to divide patients into high- and low-risk groups based on 5-year BCR outcomes (**Supplementary Table 3**), from which Kaplan-Meier (KM) curves of BCR-free survival were constructed for a subset of cases in which time-to-recurrence (BCR) data are available (**Fig. 4i**). Compared to the 2D model, the 3D model is associated with a higher hazard ratio (HR) and C-index, along with a significant *p* value (p < 0.05), suggesting superior prognostic stratification.

## Discussion

As high-resolution biomedical imaging technologies continue to evolve and generate increasingly larger datasets, computational techniques are needed to derive clinically actionable information, ideally through explainable approaches that generate new insights and hypotheses. Interpretable feature-based analysis strategies in digital pathology generally hinge upon obtaining high-quality segmentations of key structural primitives ^10,29^ (e.g., nuclei, glands, cells, collagen). However, a common bottleneck to achieving accurate segmentations is the need for large amounts of manually annotated datasets ^40^. In addition to being tedious and difficult to obtain (especially in 3D), such annotations are often performed by one or more individuals who are not representative of all pathologists, thereby introducing an early source of bias. The use of simulated data has been explored to alleviate the need for manual annotations, and has been reported to be effective for training DL-based segmentation models for highly conserved and predictable morphologies (e.g. ellipsoidal nuclei ^58,59^, or tubular vessel networks ^59,60^). However, the 3D glandular networks of prostate tissues are highly irregular and variable, making it challenging to computationally generate simulated datasets. This complex and varied 3D morphology is also in part why 2D Gleason patterns may not be ideal for characterizing prostate glands.

The ITAS3D pipeline is a general approach for the volumetric segmentation of tissue structures (e.g., vasculature / endothelial cells, neurons, collagen fibers, lymphocytes) that can be immunolabeled with high specificity and that are also discernable to a deep-learning model when labeled with small-molecule stains like our H&E analog or similar covalent stains ^61^. ITAS3D obviates the need for tedious and subjective manual annotations and, once trained, eliminates the requirement for slow/expensive antibody labeling of thick tissues (**Fig. 1**). The 2.5D segmentation approach employed in ITAS3D (i.e., image-sequence translation) offers an attractive compromise between computational speed/simplicity and accuracy for 3D objects that are relatively continuous in space (e.g., prostate glands). Details regarding 2.5D vs. 3D image translation are provided in **Supplementary Note 2**. In addition, a video summary of our ITAS3D-enabled gland-segmentation approach for PCa assessment is provided in **Supplementary Video 7**. Note that in this specific implementation of ITAS3D, intermediate images are synthetically generated to mimic an IHC stain (CK8) that is routinely used by genitourinary pathologists. Therefore, it has the added advantage of enabling intuitive troubleshooting and facilitating clinical acceptance of our computational 3D pathology approach.

In this initial clinical study, we have intentionally avoided comparing our method with extant risk classifiers or nomograms that are largely based on 2D pathology (e.g. Kattan ^62^, CAPRA ^63^, and Canary-PASS ^64^). Rather, our goal has been to demonstrate the basic feasibility and value of 3D pathology by providing a direct comparison of intuitive 3D vs. 2D glandular features. Our results show clear improvements in risk stratification based on 3D glandular features, both individually and in combination (**Fig. 4b – 4i**). As mentioned, the added prognostic value of 3D pathology is due in part to the significantly increased microscopic sampling of specimens (e.g. whole biopsies vs. sparse tissue sections). In addition, there are a number of advantages of 3D pathology datasets for computational analyses: (1) more-reliable segmentation of tissue structures due to the ability to leverage out-of-plane information (e.g. through continuity constraints); (2) the ability to quantify tissue structures more accurately in 3D while avoiding 2D artifacts ^11,65^, and (3) the ability to extract novel prognostic features that cannot be derived from 2D tissue sections (e.g. gland-skeleton features).

When trained with large numbers of images/cases and an optimal set of histomorphometric features, computational 2D pathology (based on whole slide images) has been shown to be highly prognostic ^21,66^. The goal of our study is not to suggest otherwise, but to provide early evidence of the additional prognostic value that 3D pathology can provide. The metrics presented in this study (**Fig. 4**) are intended to be comparative in nature (between 3D vs. 2D pathology) rather than regarded as definitive figures from a large prospective study.

A limitation of our implementation of ITAS3D is that the tissues used to train our deep-learning image-sequence translation modules were predominantly from Gleason pattern 3 and 4 regions, as well as benign regions. Improved ITAS3D performance over a wider range of PCa grades would be facilitated by a more-diverse set of training specimens. To more-rigorously demonstrate the ability of 3D pathology to improve upon the standard-of-care for managing PCa, larger patient cohorts and imaging datasets are needed, including prospective randomized studies on active surveillance vs. curative therapies for low-to intermediate-risk patients (e.g., the PROTECT study ^67^), as well as studies to demonstrate the ability of computational 3D pathology to predict the response of individual patients to specific treatments such as androgen deprivation and both neoadjuvant and adjuvant chemotherapy. However, as mentioned previously, the slow progression rate of most PCa cases motivates the use of archived tissues for initial validation studies, as showcased here. Finally, future studies should also aim to combine computational 3D pathology with patient metadata, such as radiomics, genomics, and electronic health records, to develop holistic decision-support algorithms ^10^.

As future work, ITAS3D can be used for the extraction and analysis of other 3D features (e.g. nuclear features, vascular features, and stromal features) to develop powerful classification models based on multiple morphologic primitives for a variety of tissue types. Annotation-free ITAS3D segmentation results, once available in sufficient quantities, can also be used to train end-to-end DL-based segmentation methods that bypass the image-translation step within ITAS3D. In the context of PCa, studies are underway to identify additional prognostic 3D features based on our unique 3D-pathology datasets. A tiered approach to analyzing PCa glandular features could be useful, such as first identifying broad classes of glandular morphologies (e.g., cribriform glands) and then analyzing class-specific features, as has been suggested in recent studies based on 2D whole slide images of PCa ^66^. Nonetheless, as an initial step towards these goals, the results of this study, as enabled by the ITAS3D computational approach, provide the strongest evidence to date in support of the value of computational 3D pathology for clinical decision support, specifically for low-to intermediate-risk PCa patients.

## Data Availability

Example prostate images for testing ITAS3D codes and models are available in a GitHub repository at https://github.com/WeisiX/ITAS3D. Full 3D prostate imaging datasets are available upon reasonable request and with the establishment of a material-transfer or data-transfer agreement. Relevant clinical data for this study are provided in **Supplementary Table 3**.

## Code availability

The Python code for the deep-learning models, and for 3D glandular segmentations based on synthetic-CK8 datasets, are available on GitHub at https://github.com/WeisiX/ITAS3D

## Acknowledgements

The authors acknowledge funding support from the Department of Defense (DoD) Prostate Cancer Research Program (PCRP) through W81XWH-18-10358 (J.T.C.L, L.D.T. and J.C.V.), W81XWH-19-1-0589 (N.P.R.), W81XWH-15-1-0558 (A.M.) and W81XWH-20-1-0851 (A.M. and J.T.C.L.). Support was also provided by the National Cancer Institute (NCI) through K99 CA240681 (A.K.G.), R01CA244170 (J.T.C.L.), U24CA199374 (A.M.), R01CA249992 (A.M.), R01CA202752 (A.M.), R01CA208236 (A.M.), R01CA216579 (A.M.), R01CA220581 (A.M.), R01CA257612 (A.M.), U01CA239055 (A.M.), U01CA248226 (A.M.), and U54CA254566 (A.M.). Additional support was provided by the National Heart, Lung and Blood Institute (NHLBI) through R01HL151277 (A.M.), the National Institute of Biomedical Imaging and Bioengineering (NIBIB) through R01EB031002 (J.T.C.L.) and R43EB028736 (A.M.), the National Institute of Mental Health through R01MH115767 (J.C.V.), the VA Merit Review Award IBX004121A from the United States Department of Veterans Affairs (A.M.), the National Science Foundation (NSF) 1934292 HDR: I-DIRSE-FW (J.T.C.L.), the NSF Graduate Research Fellowships DGE-1762114 (K.W.B.) and DGE-1762114 (L.B.), the Nancy and Buster Alvord Endowment (C.D.K), and the Prostate Cancer Foundation Young Investigator Award (N.P.R.). The training and inference of the deep learning models were facilitated by the advanced computational, storage, and networking infrastructure provided by the Hyak supercomputer system, as funded in part by the student technology fee (STF) at the University of Washington. Any opinions, findings, and conclusions or recommendations expressed in this material are those of the authors and do not necessarily reflect the views of the National Science Foundation, the National Institutes of Health, the Department of Defense, the Department of Veterans Affairs, or the United States Government.

## Author contributions

W.X., N.P.R. and J.T.C.L designed the ITAS3D segmentation pipeline. W.X. performed tri-labeling, trained the GAN models and implemented the ITAS3D segmentation. W.X. and C.K. extracted the glandular features and trained the multi-parameter model with help from P.L. and A.J. S.H. provided de-identified clinical data for our study cohort. H&E-analog staining and imaging of biopsies was performed by H.H, with help from S.K. C.M. performed thick sectioning of tissue that was used to obtain training datasets. C.M. and N.P provided guidance for the tri-labeling procedure. R.S. performed stitching and false-coloring of the OTLS whole-biopsy images. G.G. and Q.H. manually annotated 3D datasets as ground-truth for validation studies under the guidance of N.P.R. and L.D.T. A.K.G. designed and built the OTLS microscope. L.D.T. obtained tissues for this study. L.D.T., N.P.R., and J.T.C.L. designed the clinical study. J.T.C.L. supervised and coordinated the project. W.X. and J.T.C.L. led the writing of the manuscript. All authors contributed to the manuscript.

