## Supplementary materials for "Prostate cancer risk stratification via non-destructive 3D pathology with annotation-free gland segmentation and analysis"

### Supplementary Methods

Collection and processing of prostate tissue to obtain image-translation training data

Tri-labeling protocol for generating training datasets

OTLS microscopy parameters

Post-processing of training and test data

Training the image-translation models

Step 1 of ITAS3D: image-sequence translation (inference phase)

Step 2 of ITAS3D: gland segmentation based on synthetic-CK8 datasets

Benchmarking the performance of ITAS3D with alternative segmentation methods

Ground-truth annotations to validate gland-segmentation performance

Cancer-region annotations

Feature extraction: non-skeleton glandular features

Feature extraction: lumen skeleton features

**Supplementary Notes**

Omission of the “coarse-to-fine” training strategy in the vid2vid model

Comparison of redundantly processed biopsy volume when using 3D vs. 2.5D image-translation strategies

**Supplementary Figures**

Image-sequence translation model training and inference

A bivariate plot to compare the 3D SSIM metric for synthetic-CK8 images generated with and without coarse-to-fine training

Histogram of branch lengths for PCa glands

Visualization of the separation between BCR and non-BCR groups based on 3D and 2D glandular features

Use of a different fluorescent analog of H&E for model training and inference

Lumen-filling strategy

Comparing the AUC values for glandular features in cancer and benign regions

Illustration of the model-training and validation schema

**Supplementary Tables**

List of glandular features

The tri-labeling protocol.

Relevant clinical parameters for study cases (N = 50)

Comparison of 3D and 2.5D image translation in terms of the tissue volume that must be redundantly processed due to GPU limitations (which limit the 3D block sizes).

**Supplementary Videos**

Comparison of synthetic-CK8 outputs generated by models trained with and without the coarse-to-fine training strategy

Depth sequence of the benign and cancerous 3D regions shown in Fig. 3

Depth sequence of a false-colored H&E-analog dataset, a corresponding CK8-IHC dataset, and a gland-segmentation mask for a 3D prostate biopsy

Comparison between 2D image translation and 2.5D image translation

Comparison of segmentation results with ITAS3D and two baseline methods

Volume rendering of gland segmentations and skeleton networks for benign and cancerous biopsies

Video summary of ITAS3D-enabled PCa gland analysis for whole biopsies

### Supplementary References

### Supplementary Methods

**Collection and processing of prostate tissue to obtain image-translation training data.** In order to train an image-sequence translation model, we collected one FFPE block from each of nine radical prostatectomy (RP) specimens archived in an IRB-approved genitourinary biorepository at the University of Washington (UW). Based on the original pathology reports generated from the RP specimens, five specimens were from Gleason Grade Group 1 (GS = 3+3) and four specimens from Gleason Grade Group 2 or 3 (GS = 3+4 and 4+3). The imaging data from this cohort allowed us to train an image-translation model for low- to intermediate-risk PCa (Grade Group 1-3).

For deparaffinization, the FFPE blocks were first heated at 75°C for 1 hour until the outer paraffin wax was melted. The tissue blocks were then treated 2× with 500 ml of 75°C xylene for 24 hours. A hotplate with magnetic stirrer was used to maintain the temperature of the xylene and to promote fluid convection around the specimens. Next, we used a vibratome to cut a 200-μm-thick slice from the surface of each tissue block. This thickness was optimized to balance two factors: 1) providing sufficient 3D context to train and test the image-sequence translation model and 2) allowing for uniform antibody diffusion and staining within the tissue slices. The tissue slices averaged about 1.5 cm × 1 cm in their lateral dimension. Prior to staining, each tissue slice was cut into smaller pieces, measuring approximately 0.5 cm × 0.5 cm × 200 μm, to further promote fluid convection around all specimens during the staining protocol.

**Tri-labeling protocol for generating training datasets.** We performed a tri-labeling protocol (see **Supplementary Table 2**) that utilized SYTO™ 85 (Cat: S11366, ThermoFisher) as a nuclear stain (hematoxylin analog), Alexa Fluor™ 488 NHS Ester (Cat: A20000, ThermoFisher) as a cytoplasmic stain (eosin analog) [61], and a CK8-targeted monoclonal antibody (Cat: MA5-14088,

ThermoFisher) for immunofluorescence labeling. Our tri-labeling protocol was adapted from the iDISCO protocol [14]. We first labeled the tissue with an Alexa Fluor™ 488 NHS ester (binds to all proteins) prior to performing CK8 immunostaining in order to prevent the NHS ester from staining the CK8 antibody. This ensures unbiased training of the H&E-to-CK8 translation model. CK8 immunostaining included a primary and secondary antibody staining step. Nuclear staining with SYTO™ 85 was performed afterwards, followed by tissue dehydration in ethanol and optical clearing (index-matching, n = 1.56) with ethyl cinnamate (Cat: 112372, Sigma-Aldrich).

The fluorescent analog of H&E used for generating training datasets (i.e., SYTO™ 85 + Alexa Fluor™ 488 NHS Ester, "S&N") was slightly different from the H&E analog used for whole-biopsy staining in our inference datasets (i.e. TO-PRO-3 + eosin, "T&E"). The S&N version of our H&E analog was used for generating the training data so that the CK8 immunofluorescence could be placed in the longest-wavelength channel. This was done to ensure that there was negligible crosstalk of the CK8 immunofluorescence into the H&E-analog wavelength channels (for unbiased training). Despite this difference in the H&E-analog staining protocols, our trained model was shown to be applicable to both S&N- and T&E-labeled tissues as inputs. High-fidelity image translation was achieved in both cases with minimal differences in appearance (**Supplementary Fig. 5**).

**OTLS microscopy parameters.** For imaging the 200-μm thick tri-labeled prostate tissue sections (training specimens), the Alexa Fluor™ 488 NHS Ester was excited at 488 nm and imaged through an emission bandpass filter (FF03–525/ 50–25, Semrock). The nucleic-acid-targeting fluorophore, SYTO™ 85, was excited at 561 nm and imaged through another bandpass filter (FF01–618/50–25, Semrock). CK8 immunofluorescence (Alexa Fluor™ 647 conjugated secondary antibody) was excited at 638 nm and imaged through a third bandpass filter (FF01–721/ 65–25, Semrock). Raw OTLS images were downsampled by 2× in all three dimensions (which resulted in a sampling pitch of ~0.9 μm/pixel) and fused into a continuous 3D dataset using the BigStitcher plug-in [68] in ImageJ. This was done to alleviate memory requirements and to accelerate image-translation computations. However, this level of downsampling still preserved sufficient sub-cellular details for effective image translation and visual inspection by pathologists. From the fused 3D imaging data, we extracted image volumes measuring 1024 × 1024 × 50 pixels, which were treated as image sequences with 50 depth levels (corresponding to ~45 μm in depth). These image sub-blocks were used for training and testing of the image-translation model.

For imaging the T&E-labeled prostate biopsies, eosin was excited at 488 nm and imaged through an emission bandpass filter (FF03–525/ 50–25, Semrock). The nucleic-acid-targeted fluorophore, To-PRO™-3 Iodide, was excited at 638 nm laser and imaged through another bandpass filter (FF01–721/ 65–25, Semrock). The fused datasets were downsampled by 2× in all three dimensions using the BigStitcher plug-in [68] in ImageJ.

**Post-processing of training and test data.** The small-molecule H&E-analog stain was highly reproducible and generated spatially uniform images [13,19]. Therefore, data screening/cleaning was primarily based on the image quality of the CK8 immunofluorescence channel. In the training and testing datasets, we only included tissue regions with spatially uniform CK8 labeling (based on visual inspection). To further enhance the contrast and uniformity of the CK8 training data (and hence the uniformity of the image-translation outputs), contrast-limited adaptive histogram equalization (CLAHE) [69] and flat-fielding [70] were applied on the CK8 datasets. For the training set, we obtained a total of 1,806 tri-labeled image sequences (each containing 50 levels) as described in the last section. For the validation set, which was used to calculate performance metrics, we set aside 58 image sequences that were not used in the training phase. The H&E-analog channels were used as model inputs, and corresponding co-registered CK8 images was used as model targets (desired outputs) for our GAN-based supervised training strategy.

**Training the image-translation models.** Generative adversarial networks (GANs) [71] provide a generalizable model and loss function to solve different image-translation problems that traditionally would require the design of specialized models and loss functions by practitioners with extensive field-specific expertise. In GANs, the discriminator's goal is to classify images as real or fake (generated), while the generator is trained to synthesize realistic output data that can "fool" the discriminator. As an extension of the GAN structure, conditional GANs (cGANs) are structured such that both the generator and discriminator are conditioned with prior information (in our case, H&E-analog inputs), such that the model can learn how to map inputs (H&E analog) to outputs (synthetic CK8) in a supervised manner (**Supplementary Fig. 1a**).

We adapted *pix2pix* [72], a prior implementation of cGANs for single-level 2D image translation, in order to initialize our image-sequence translation workflow. The *pix2pix* framework directly conditions the generator ($G_{imge}$ in **Supplementary Fig. 1a**) with input images that are paired with target images. Our adapted generator uses a U-Net [52] architecture with 9 layers of downsampling and up-sampling, and the discriminator ($D_{image}$ in **Supplementary Fig. 1a**) uses a convolutional PatchGAN [73] classifier. The *pix2pix* model was first introduced for tasks such as label-to-photo or pose-to-photo translations but has also been adapted for biomedical image translations in many recent studies [45,74]. For *pix2pix* training, from each 50-level image sequence, we selected five levels of 2D images that were 10 pixels apart (~9 µm) to maximize diversity in the training set. This yielded 9030 images in total for *pix2pix* training, each of which was 1024 × 1024 pixels in size. Each 2D image level was treated as an independent input during training. Hyperparameters were set as default values (identical to the original *pix2pix* model) except for those specified in the training script provided in our GitHub repository. The training required 200 epochs for 44 hours with a 12-GB NVIDIA Tesla P100 GPU on a standard node of the UW Hyak high-performance computation (HPC) cluster.

For “2.5D” image-sequence translation, we adapted a video-translation model, *vid2vid* [47], which performs image conversion frame-by-frame (originally in a “time stack”) and utilizes information in adjacent frames to ensure that temporal continuity is maintained. Given that we treat a 3D image as a “z-stack” of 2D images, we can apply this same concept to improve continuity in the depth dimension. As shown in **Supplementary Fig. 1b**, the sequence generator ($G_{sequence}$) synthesizes CK8 images in a level-by-level manner by taking into account the synthetic CK8 images generated at two previous levels along with the corresponding H&E analog images at each level (i.e., the current plus two previous levels). Similar to *pix2pix*, a multi-scale PatchGAN architecture is adopted for the image discriminator ($D_{image}$). In addition, a multi-scale sequence discriminator ($D_{sequence}$) is trained to ensure small- and large-scale consistency in the 3D spatial domain. All 1,806 image sequences (50 levels per sequence) were used for the *vid2vid* training set.

**Step 1 of ITAS3D: image-sequence translation (inference phase).** Whole-biopsy H&E-analog datasets are first sub-divided into blocks of size 1024 × 1024 × 712 pixels with 25% overlap along the biopsy-axis direction (the long axis of the cylindrical biopsy). For level-by-level (2.5D) inference of synthetic CK8 images, each depth level takes into account the synthetic CK8 images generated at two previous levels. Therefore, in order to initiate this process, a 2D translation model (the *pix2pix* model in our case) is required to generate the first two synthetic images within the depth sequence (**Supplementary Fig. 1c**). For each image-sequence block, we start the image-sequence translation bi-directionally from the center of the block. This ensures that the initial images are of high-quality and that we can avoid irregularities at the tissue edges (e.g., staining artifacts and surface irregularities at the edges of the biopsies). We split each block into two halves with 34 overlapping levels (**Supplementary Fig. 1c**). As mentioned, to initialize the image-sequence translation process, 2D image translation (*pix2pix*) is first performed on the bottom two levels of the top half of the block, and on the top two levels of the bottom half of the block. The image translation of each biopsy (usually consisting of 10-15 blocks with 25% overlap between adjacent blocks) takes ~6 hours with a 12-GB NVIDIA Tesla P100 GPU on a standard node of the UW Hyak HPC cluster. Hyperparameters are set as default values (identical to the original *vid2vid* model) except for those specified in the inference script provided in our GitHub repository. Once image-sequence translation is completed for both halves, the two halves are merged: the first 12 levels in each half are discarded (i.e., they are only used to initiate the image-sequence translation processes), and the remaining 10 overlapping levels are linearly blended (the intensities are smoothly adjusted between the two images so that one image fades out as the other image transitions in). Finally, the synthetic-CK8 image blocks are mosaicked with linear blending using the ImageJ “stitching” plug-in [75].

**Step 2 of ITAS3D: gland segmentation based on synthetic-CK8 datasets.** Prior to performing gland segmentation, the synthetic-CK8 images are downsampled by 2× in all 3 dimensions (1.8 µm/pixel) to reduce memory requirements and computational times while still enabling gland segmentations to be performed at a reasonable spatial resolution. The resulting synthetic-CK8 datasets for each biopsy are approximately 7000 (length) × 512 (width) x 356 (depth) pixels in size. First, to enhance the contrast of the boundary of the epithelium with respect to the background, a 3×3 edge-sharpening filter is applied to each sagittal 2D level within the 3D dataset (the sagittal view is the typical *en face* view of a biopsy seen by pathologists). Then, a preliminary segmentation mask for the epithelium is obtained via Otsu thresholding. The CK8 biomarker is expressed in the cytoplasm of the luminal epithelial cells. Therefore, in order to fill in the unstained nuclei and to acquire a solid epithelium mask, a binary closing routine (dilation followed by erosion) is performed using a 3×3 structural element with a square connectivity equal to one, followed by a small-hole filling routine applied to the epithelium mask.

To segment the lumen, we first fill in any 2D contours enclosed by the epithelium mask in a level-by-level manner along 3 orthogonal directions (see **Supplementary Fig. 6**), which are then combined. This allows most lumen spaces to be filled in accurately even in the presence of small and sparse gaps in the 3D epithelium mask that may arise due to imperfections in the synthetic-CK8 images. However, this method occasionally introduces false-positive lumen regions that actually correspond to cytoplasm regions. A cytoplasm mask, obtained with Otsu thresholding of the eosin channel, is used to remove these false-lumen regions. We finalize the semantic segmentation of glands by taking the union of the lumen and epithelium masks with some minor adjustments, such as noise removal and filling in of small holes. In addition, a stroma mask is obtained by applying an active contour algorithm [48] to the cytoplasm channel. For each biopsy, the computational time for gland segmentation when starting from a synthetic CK8 dataset is ~30 minutes for a 3.20GHz CPU (Intel® Xeon® Gold 6134) with 64Gb of RAM.

**Benchmarking the performance of ITAS3D with alternative segmentation methods.** The effectiveness of ITAS3D was compared with two well-known object-segmentation methods. The first benchmarking method utilized traditional 3D image-processing techniques, starting with a watershed method [51] extended to 3D, which was applied on the eosin-analog channel to identify candidate lumen regions only (segmenting the epithelial cells was not successful with this method). Here, the 3D-watershed algorithm was initiated at marker points that were identified with an Otsu thresholding routine applied on the same eosin-analog images. Likewise, nuclei were detected by applying another watershed-based segmentation method on the hematoxylin-analog images [76]. Candidate lumen regions in which the majority of the boundary pixels were not adjacent (within 10 μm) to segmented nuclei were eliminated due to the fact that true lumen regions are always enclosed by epithelial cells. The second benchmarking method was a convolutional neural network, U-Net [52], originally developed for biomedical image segmentation. To train the 2D U-Net model, patches were extracted from 15 ROIs from five biopsies (3 ROIs from each). Manual ground-truth annotations of glands were performed under the guidance of a board-certified pathologist (N.P.R.). Four biopsies were used for training with the remaining biopsy used to validate the accuracy of the model. For each 512 × 512 patch, a split ratio of 4:1 was utilized for training vs. validation [52,77,78], which resulted in 1600 patches for training, and 400 for validation. During training, the patches were further augmented via random left or right flipping manipulations, and 90° rotations, to increase the robustness of the model.

**Ground-truth annotations to validate gland-segmentation performance.** Ten 0.2-mm$^3$ regions (512 × 512 × 100 voxel each) from different patients were randomly selected from the testing dataset (tri-labeled 3D images not used for training). These regions were manually annotated based on H&E-analog and real-CK8 immunofluorescence images to obtain a ground-truth set of 3D gland segmentations (for both the lumen mask and epithelium mask). These manual annotations were performed using a commercial image-analysis software package, Aivia (Leica Microsystems), under the guidance of a board-certified genitourinary pathologist (N.P.R.).

**Cancer-region annotations.** Cancer-enriched regions in each biopsy were annotated by recording the coordinates of those regions along the axis of each cylindrically shaped biopsy. This was performed based on a level-by-level visual inspection of the 3D biopsy datasets under the guidance of a board-certified genitourinary pathologist (N.P.R.). We analyzed the glandular features only within the cancerous regions of each biopsy. This was motivated by our finding that histomorphometric features from cancer glands exhibit higher correlation with BCR outcomes than those from adjacent benign glands (**Supplementary Fig. 7)**. However, due to the fact that our cancer-region annotations are only performed along the long axis of each cylindrical biopsy (i.e., rough annotations), as well as the observation that benign and cancer glands can be intermixed within certain tissue regions, a more-precise and accurate method to identify individual cancer glands would likely further improve the prognostic value of our methods in the future.

**Feature extraction: non-skeleton glandular features.** From the 3D gland segmentations, 12 non-skeleton glandular features relating to gland size and shape were computed. These features are presented in **Supplementary Table 1** along with a brief description and AUC value for each feature. In order to explore the advantages of extending 2D glandular features into 3D, we analyzed features that have direct 2D vs. 3D analogs. The first feature set characterizes the volumetric extent of different tissue compartments (lumen, epithelium, and stroma). The other feature sets characterize the shape of the different compartments, which are derived from a "triangle mesh" that approximates the compartments [79]. Such features include the surface-area-to-volume ratio of an object (in 3D) or circumference-to-area ratio (in 2D); the average surface curvature (in 3D) or boundary curvature (in 2D) of an object [35]; and the gland-to-convex-hull ratio (G/H), defined as the volume ratio (in 3D) or area ratio (in 2D) of the gland mask (epithelium + lumen) to the convex hull that circumscribes the gland. The non-skeleton glandular features were extracted using the "regionprops", "mesh_surface_area", and "ConvexHull" methods from Python's "scikit-image" and "scipy" packages. 3D curvature values were calculated with the "discrete_gaussian_curvature_measure" method from the "trimesh" library using meshes generated by the "marching_cubes" method in "scikit-image".

**Feature extraction: lumen skeleton features.** Extraction of the lumen skeleton (centerline of the lumen) was based on a 3D-thinning algorithm [80]. As mentioned in the main manuscript, meaningful 2D analogs of the 3D skeleton-based features do not exist (**Supplementary Table 1**). Before the skeleton extraction, we down-sampled the segmentation mask by 4× and applied a binary erosion (with a spherical kernel of a 2-voxel radius) to prevent false branches from being introduced due to fine surface irregularities. For the skeleton networks, a junction is defined as the point where a lumen splits into two or more lumens. A branch is defined either as a segment between a junction and a junction, a junction and an end point, an end point to an end point, or an isolated loop [81]. We extracted 5 different skeleton features: mean branch length, standard deviation of the branch lengths, median tortuosity (defined as the ratio of branch length to the Euclidean distance between the two end points of the branch), standard deviation of the tortuosity, and branch connectivity. Here, branch connectivity is defined as the ratio of the total number of branches to the total number of connected sets of branches within each biopsy.

### Supplementary Notes

#### Supplementary Note 1 | Omission of the “coarse-to-fine” training strategy in the vid2vid model

The coarse-to-fine strategy [1] was a technique employed within the original *vid2vid* model to generate high-resolution videos. In this method, the resolution of the training images was increased incrementally (2× at a time, starting from downsampled images) while adding resolution-matched residual blocks [2] to the front and back of the generator during the training process until the desired target resolution was reached. With the coarse-to-fine training strategy, image-sequence translation training was performed over 54 epochs, which took 2 months. Without the coarse-to-fine training strategy, only 20 epochs were required, taking about 1 month, but with negligible performance loss (see **Supplementary note 1**, **Supplementary Fig. 2** and **Supplementary Video 1**). Both models were trained with a 12-GB NVIDIA Tesla P100 GPU on a standard node at the UW Hyak HPC cluster. Note that if the resources are available, the training can be done with multiple GPUs (e.g., 8 GPUs in the *vid2vid* paper) to potentially shorten the training time to a few days. Hyperparameters were set as default values (identical to the original *vid2vid* model) except for those specified in the training script provided in our GitHub repository.

The “coarse-to-fine” training strategy was initially proposed to improve the translation of real-world inputs, such as natural-scenery videos, over a large range of spatial scales [1]. Our speculation is that unlike such videos, our CK8 image translation operates on a more-limited range of micro-scale structures (i.e., close-packed arrangements of luminal epithelial cells surrounding the prostate glands). Therefore, the “coarse-to-fine” method, which significantly increases the training time, does not noticeably improve the model’s performance.

**Supplementary Note 2 | Comparison of redundantly processed biopsy volume when using 3D vs. 2.5D image-translation strategies**

There are practical advantages for our 2.5D image-translation-based segmentation approach (ITAS3D) in comparison to an approach that operates on whole 3D data cubes. For computational analyses, the maximum allowable size of a 3D data cube (volumetric sub region) is typically limited by GPU memory. For instance, with the 3D image-translation method, *vox2vox* [3], which uses a 3D U-Net [4] structure as its generator, a 3D block size that pushes against the computational limits of a NVIDIA GeForce RTX 2080 Ti GPU during the inference phase is ~ 160 × 190 × 130 pixels. With the OTLS microscope used in this study, this corresponds to a physical tissue volume of about 140 × 170 × 110 $\mu m^3$. This small block size, which results in many prostate glands being truncated, could lead to errors (e.g., edge and stitching errors) when performing 3D gland segmentations. A common strategy to reduce stitching errors is to include a larger amount of overlap between neighboring sub-volumes, but this would further increase computational times.

As a rough estimate based on our experiences with ITAS3D and published results for *vox2vox* [3] (see **Supplementary Table 4**), and assuming a 25% overlap ratio between image blocks processed with a 12-GB GPU, approximately 30% of the tissue volume in each biopsy would be redundantly processed using a 2.5D image-translation method (e.g. ITAS3D), whereas more than 100% of each biopsy volume would be redundantly processed using 3D image translation (*vox2vox*) due to the overlap between sub-volumes. This would contribute to increased computational times for a 3D image-translation strategy.

### Supplementary Figures

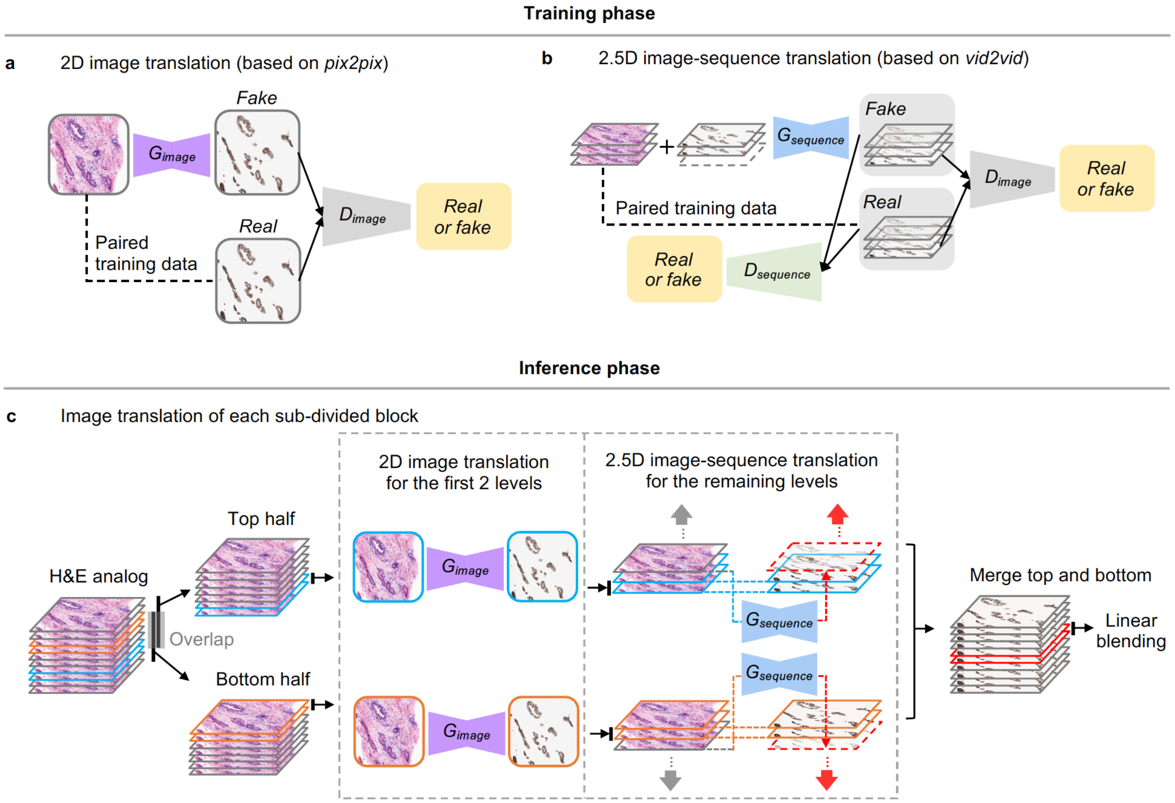


**Supplementary Fig. 1 | Image-sequence translation model training and inference. a**, Image translation in this study is based on previously developed conditional GANs (*pix2pix* for 2D image translation, and *vid2vid* for 2.5D image translation), which are trained with paired imaging data in a fully supervised manner. For 2D image translation, the generator ($G_{image}$) is trained to translate 2D H&E-analog images into 2D synthetic-CK8 images that cannot be distinguished from real CK8 images by the discriminator ($D_{image}$), which is adversarially trained to classify between real and fake (synthetic) CK8 images. **b,** For 2.5D image translation, a 3D image is regarded as a sequence of 2D images. The generator ($G_{sequence}$) is trained to produce synthetic CK8 images in a level-by-level manner, conditioned with both an H&E-analog input image at each level, as well as H&E-analog and synthetic-CK8 images at two previous levels to ensure spatial continuity between adjacent images/levels within the sequence. Meanwhile, an image discriminator ($D_{image}$) and a sequence discriminator ($D_{sequence}$) are adversarially trained to classify between real or fake (synthetic) 2D images and image sequences, respectively. **c**, In the inference phase, to ensure robust image translation, each H&E-analog image block is split into a top and bottom half with overlapping regions. This allows image-sequence translation to be initiated from the center of the biopsy, where image-quality is optimal and tissue-edge artifacts are avoided. For each half, image-sequence translation is initiated with 2D image translation of the first two levels (in blue for the top half, in orange for the bottom half), which transitions to 2.5D image-sequence translation for the remaining levels. The final synthetic-CK8 dataset is obtained by merging the top and bottom halves of the block with linear blending of the overlapping middle levels (in red).

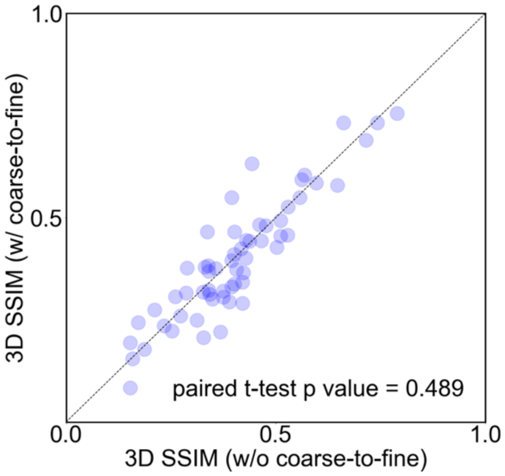


**Supplementary Fig. 2 | A bivariate plot to compare the 3D SSIM metric for synthetic-CK8 images generated with and without coarse-to-fine training.** The testing dataset contains 58 tissue volumes, each with a size of ~0.2-mm$^3$ (1024 × 1024 × 200 pixel). The *p* values (two-sided paired t-test) show no significant differences between the SSIM distributions for synthetic-CK8 images generated by the two models (with and without the coarse-to-fine training strategy).

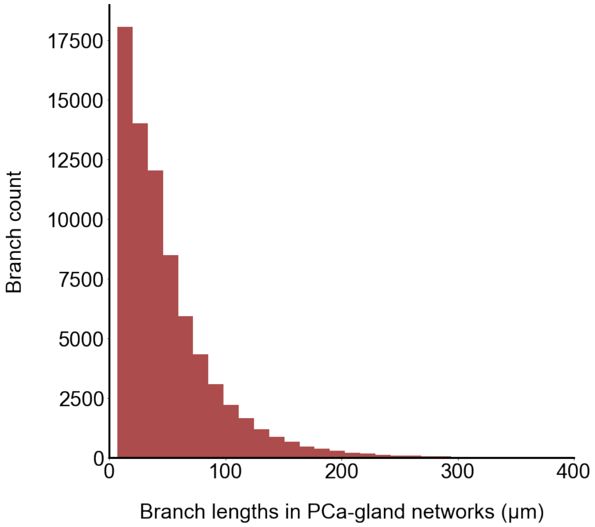


**Supplementary Fig. 3 | Histogram of branch lengths for PCa glands.** The diameter of the biopsies (1-mm) imaged in this study is large enough to accurately quantify the majority of branch lengths for PCa glands.

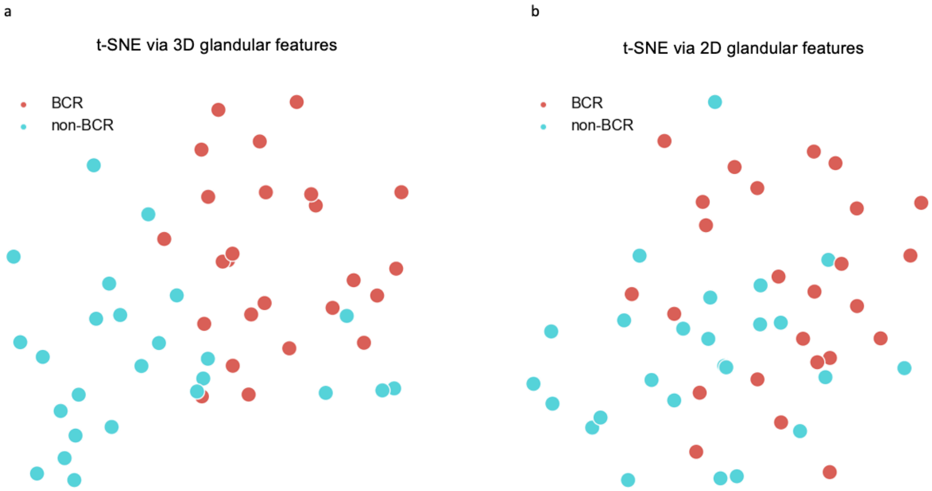


**Supplementary Fig. 4 | Visualization of the separation between BCR and non-BCR groups based on 3D and 2D glandular features.** The 25 BCR cases and 25 non-BCR cases were mapped to a 2-dimensional space using *t*-SNE. The separation between the two groups is more evident when the *t*-SNE analysis is based on 3D (**a**) rather than 2D (**b**) glandular features.

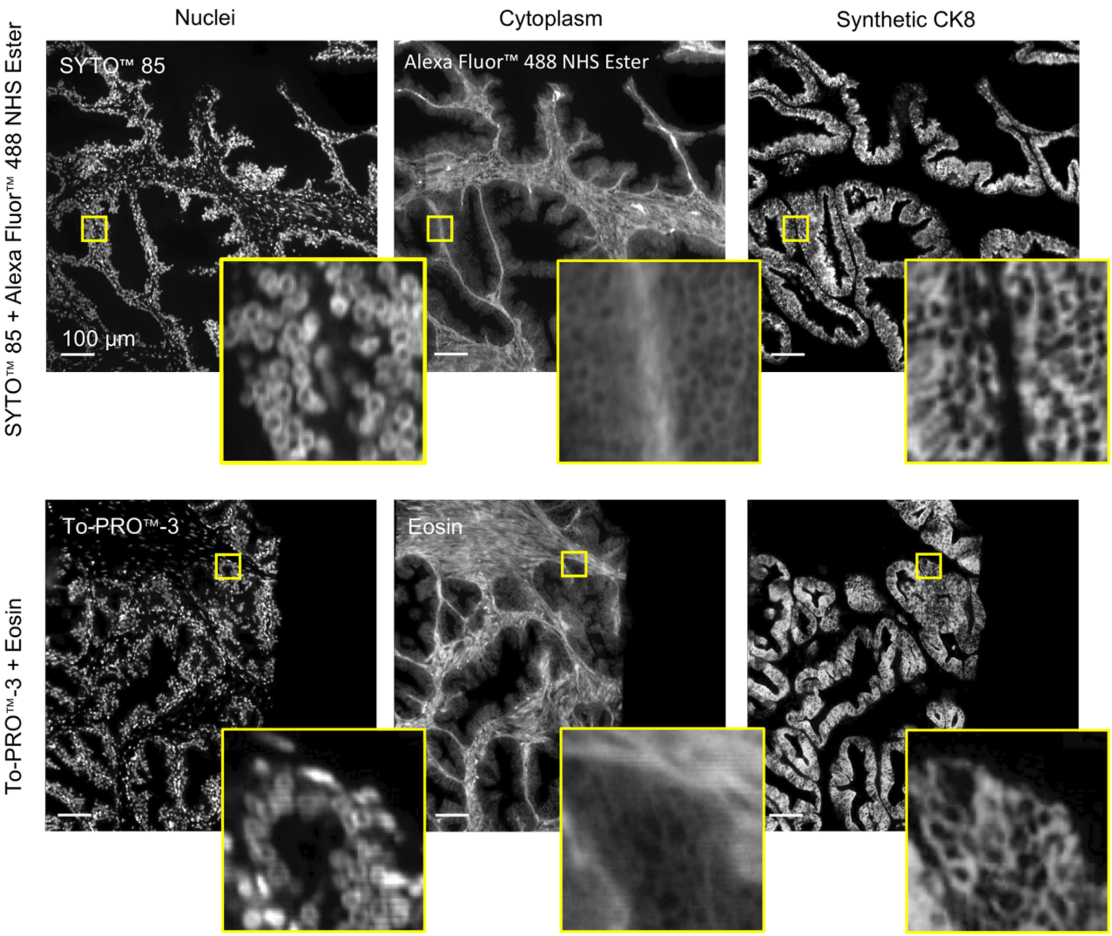


**Supplementary Fig. 5 | Use of a different fluorescent analog of H&E for model training and inference.** Although our image-sequence translation model is trained with a H&E analog that is slightly different from the H&E analog used for our inference datasets, the trained model performs comparably well in both cases. The top and bottom rows show image-translation results based on tissues stained with an "S&N" protocol (used for training) versus a "T&E" protocol (used in our clinical studies). Minimal differences are seen in the synthetic-CK8 output images (far right). This is likely due to the highly similar appearance of tissue stained with the two H&E-analog protocols (as shown). Scale bar: 100 µm.

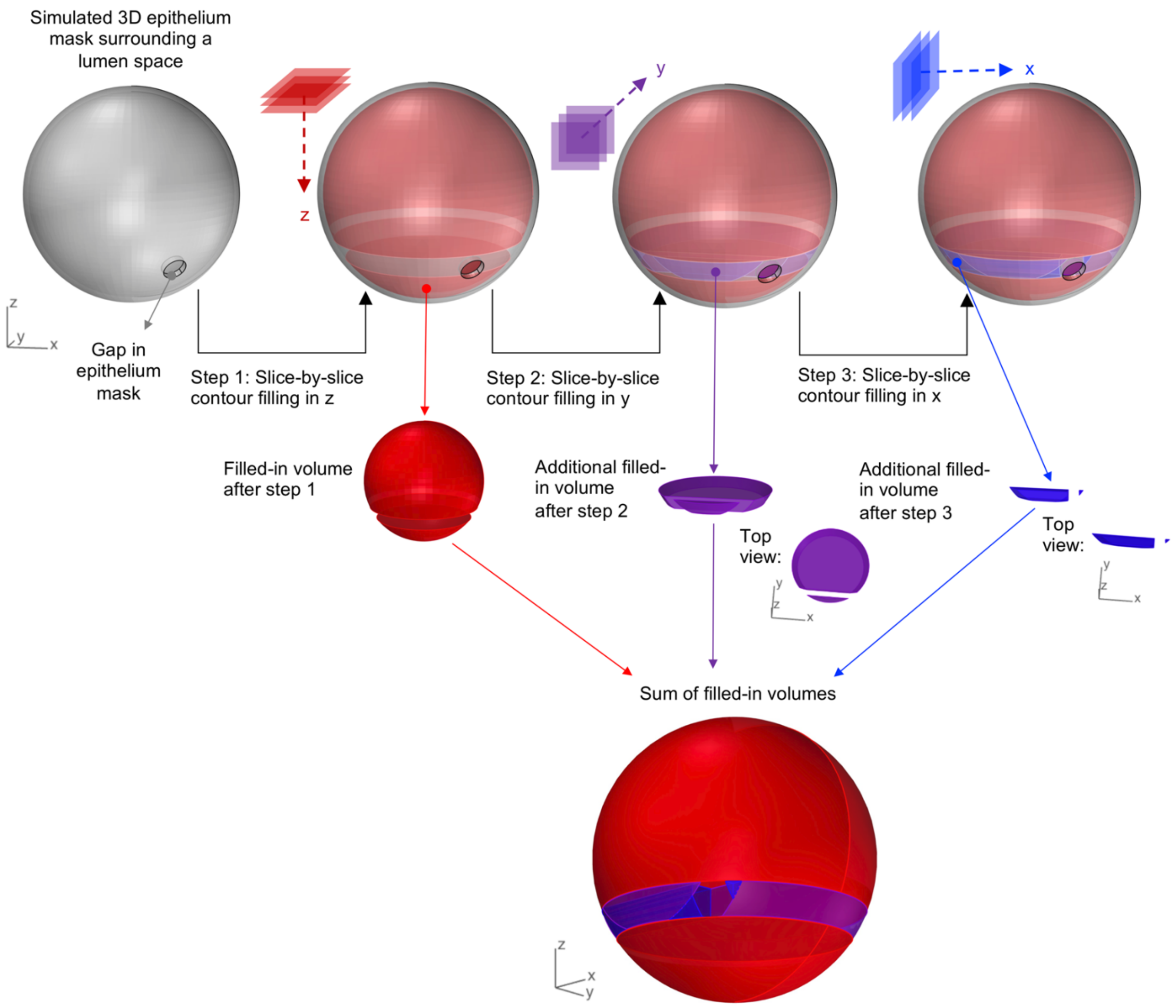


**Supplementary Fig. 6 | Lumen-filling strategy.** Epithelial cells should ideally completely enclose all lumen regions. However, due to imperfect labeling, sparse and small gaps in the epithelia occasionally appear. This leads to errors when attempting to identify the lumen spaces with a slice-by-slice contour-filling routine (as shown in step 1). However, by performing slice-by-slice contour filling along 3 orthogonal directions and combining the results, most lumen spaces are accurately filled in. We have found this method to be superior to standard 3D methods for filling in enclosed surfaces.

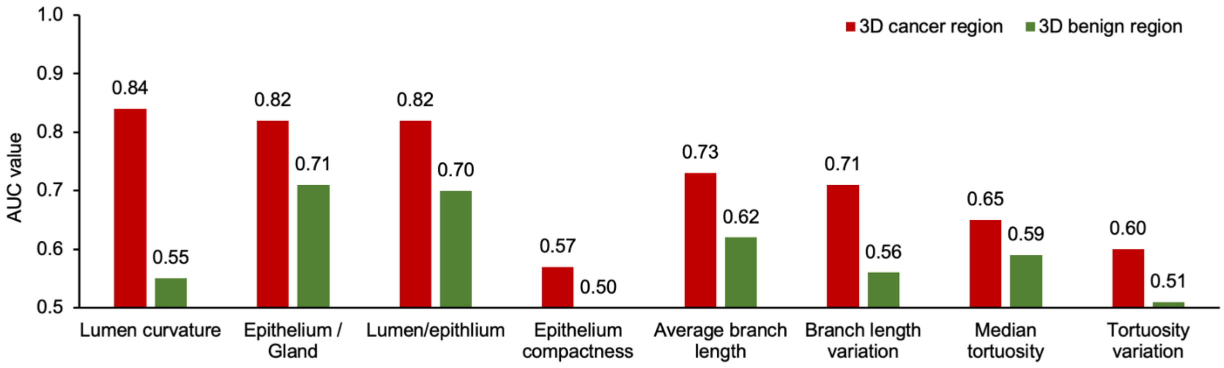


**Supplementary Fig. 7 | Comparing the AUC values for glandular features in cancer and benign regions.** Patient-level AUC values for glandular features (4 non-skeleton and 4 skeleton-based features are shown here) are higher for glands in cancerous regions than in adjacent benign regions. We therefore focused our analyses on cancer-enriched regions of each biopsy.

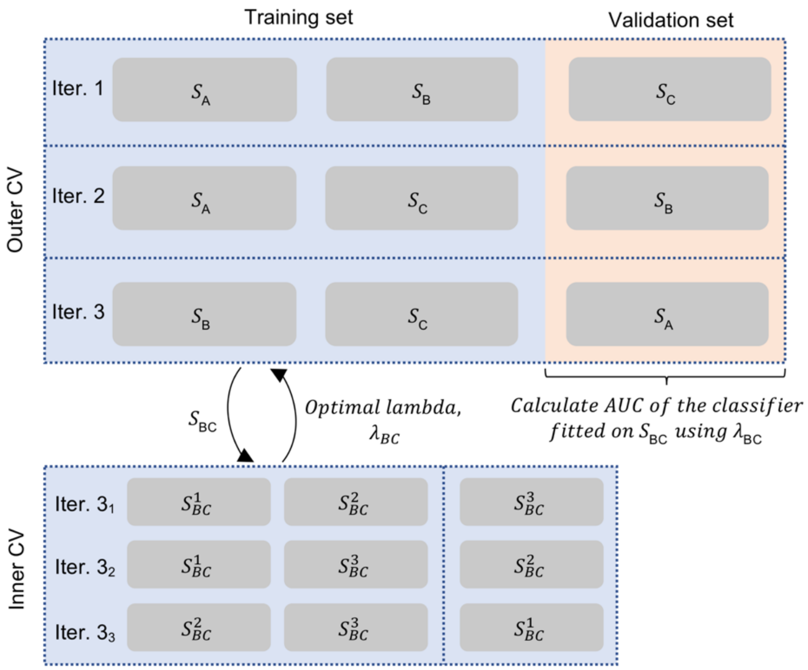


**Supplementary Fig. 8 | Illustration of the model-training and validation schema.** A nested 3-fold cross validation (CV) was used for model training and validation. The inner CV was performed at each iteration of the outer CV to determine the optimal model parameter, λ. In the outer CV, the model was developed based on the training fold (2/3 of the cases) using the optimal λ value. Model performance metrics were then quantified based on the validation fold (remaining 1/3 of cases) to calculate the AUC. The nested CV ensures that there is no overlap between the data used to develop the classification model and the data used to evaluate the performance of the model. The nested CV was performed 200 times, generating 600 AUCs (3 AUCs from each iteration). The average and standard deviation of the AUCs were calculated and compared.

### Supplementary Tables

#### Supplementary Table 1 | List of glandular features

| Feature category | Feature set | 3D glandular features (*G: gland* [1], *E: epithelium, L: lumen, S: stroma)* | 3D AUC value | 2D glandular features | 2D AUC value |
|---|---|---|---|---|---|
| Non-skeleton features | Size | *Volume (L) / volume (E)* | 0.82 | *Area (L) / area (E)* | 0.79 |
| | | *Volume (E) / volume (G)* | 0.82 | *Area (E) / area (G)* | 0.78 |
| | | *Volume (S) / volume (G)* | 0.67 | *Area (S) / area (G)* | 0.68 |
| | Compactness | *Surface area (G) / volume (G)* | 0.68 | *Circumference (G) / area (G)* | 0.63 |
| | | *Surface area (E) / volume (E)* | 0.57 | *Circumference (E) / area (E)* | 0.55 |
| | | *Surface area (L) / volume (L)* | 0.69 | *Circumference (L) / area (L)* | 0.75 |
| | Irregularity | *Volume (G) / convex hull volume (G)* | 0.72 | *Area (G) / convex hull area (G)* | 0.62 |
| | | *Volume (E) / convex hull volume (E)* | 0.52 | *Area (E) / convex hull area (E)* | 0.72 |
| | | *Volume (L) / convex hull volume (L)* | 0.67 | *Area (L) / convex hull area (L)* | 0.51 |
| | Boundary Curvature | Average Gaussian curvature (absolute values) of the *G* surface | 0.79 | Average curvature of the *G* boundary [5]. | 0.72 |
| | | Average Gaussian curvature (absolute values) of the E surface | 0.80 | Average curvature of the *E* boundary [5]. | 0.64 |
| | | Average Gaussian curvature (absolute values) of the *L* surface | 0.84 | Average curvature of the *L* boundary [5]. | 0.74 |
| Skeleton features | Lumen skeleton | Average branch length | 0.73 | N /A | |
| | | Standard deviation of the branch length | 0.71 | | |
| | | Median tortuosity [2] for all branches | 0.65 | | |
| | | Standard deviation of the tortuosity for all branches | 0.6 | | |
| | | Branch connectivity [3] | 0.58 | | |

[1] G = E + L
[2] Tortuosity is defined as: branch length / Euclidian distance between two end points
[3] Branch connectivity is defined as: total number of branches / total number of connected sets of branches

### Supplementary Table 2 | The tri-labeling protocol

| Time | Steps |
|---|---|
| **Week 1** | |
| Day 2 | Wash samples with PBS (1× phosphate-buffered saline) for 1h, then in 25% methanol (in PBS) for 1h, 50% methanol for 1h, 75% methanol for 1h, and 100% methanol for 1h at room temperature. Samples are then put into 100% methanol for storage. |
| Day 3 | Chill samples at 4°C for 1h, then bleach in 5%$H_2O_2$ in methanol at 4°C overnight. |
| Day 4 | Wash samples in 75% methanol (in PBS) for 1h, then in 50% methanol for 1h, 25% methanol for 1h, 100% PBS for 1hr at room temperature. For cytoplasm staining, samples are incubated in 5μg/ml Alexa Fluor™ 488 NHS ester (dissolved in pH 5 PBS, where the pH is adjusted with HCl, Cat: A20000, ThermoFisher) at 37°C overnight. |
| Day 5 | Wash samples in PBS/0.2% Triton X-100 for 1h at room temperature twice. Samples are kept in PBS/0.2% Triton X-100 at room temperature. |
| **Week 2** | |
| Day 1 | Samples are incubated in PBS/0.2% Triton X-100/20% DMSO/0.3M glycine, at 37°C overnight. |
| Day 2 | Block samples with PBS/0.2% TritonX-100/10% DMSO/6% Donkey Serum/3mM $NaN_3$, at 37°C overnight. |
| Day 3 | Samples are incubated with the primary antibody (Cytokeratin 8/18 Monoclonal Antibody 5D3, 1:20 diluted, Cat: MA5-14088, ThermoFisher) in PBS/0.2%Tween-20/10μg/ml heparin (PTwH)/5%DMSO/3% Donkey Serum/3mM $NaN_3$ at 37°C until the next step. |
| **Week 3** | |
| Day 2 | Wash samples in PTwH for 15 min, 30 min, 1h and 1h at room temperature. Samples are then kept in PTwH overnight at room temperature. |
| Day 3 | Samples are incubated with the secondary antibody (Alexa Fluor 647 AffiniPure Donkey Anti-Mouse IgG, 1:100 diluted, Cat: 715-605-150, JacksonImmunoResearch) in PTwH/3% Donkey Serum/3mM $NaN_3$ at 37°C. |
| **Week 4** | |
| Day 1 | Samples are incubated in PTwH for 15 min, 30 min, 1h and 1h at room temperature. For nuclear staining, samples are incubated 5uM SYTO™ 85 (in PTwH, Cat: S11366, ThermoFisher) overnight at 37°C. |
| Day 2 | Wash samples with PBS for 1h, then incubate with 25% ethanol (in PBS) for 1h, 50% ethanol for 1h, 75% ethanol for 1h, and 100% ethanol for 1h at room temperature. Samples are then kept in 100% ethanol overnight at room temperature. |
| Day 3 | For optical clearing, samples are incubated in ethyl cinnamate for 30 min 2x at room temperature. Samples are then ready for 3D OTLS microscopy. |

### Supplementary Table 3 | Relevant clinical parameters for study cases (N = 50)

| Case # | Eligibility | BCR category [1] | Age at diagnosis | Days to recurrence post-RP | Multiparameter model classification based on 2D features [2] | Multiparameter model classification based on 3D (w/o skeleton) features [2] |
|---|---|---|---|---|---|---|
| 1 | Non-recurrent | 0 | 50-54 | N/A | Low risk | Low risk |
| 2 | Recurrent (<5 years) | 1 | 65-69 | 669 | Low risk | High risk |
| 3 | Recurrent (<5 years) | 1 | 55-59 | Data not available | Low risk | Low risk |
| 4 | Recurrent (<5 years) | 1 | 55-59 | Data not available | High risk | High risk |
| 5 | Non-recurrent | 0 | 60-64 | N/A | High risk | High risk |
| 6 | Recurrent (<5 years) | 1 | 70-74 | Data not available | High risk | High risk |
| 7 | Recurrent (>5 years) | 0 | 45-49 | Data not available | Low risk | Low risk |
| 8 | Non-recurrent | 0 | 55-59 | N/A | Low risk | Low risk |
| 9 | Recurrent (<5 years) | 1 | 70-74 | Data not available | High risk | High risk |
| 10 | Recurrent (<5 years) | 1 | 45-49 | Data not available | High risk | Low risk |
| 11 | Recurrent (<5 years) | 1 | 55-59 | 1491 | Low risk | High risk |
| 12 | Non-recurrent | 0 | 60-64 | N/A | Low risk | Low risk |
| 13 | Non-recurrent | 0 | 55-59 | N/A | High risk | Low risk |
| 14 | Non-recurrent | 0 | 55-59 | N/A | Low risk | Low risk |
| 15 | Recurrent (>5 years) | 0 | 60-64 | 1949 | High risk | High risk |
| 16 | Recurrent (<5 years) | 1 | 50-54 | Data not available | High risk | High risk |
| 17 | Non-recurrent | 0 | 50-54 | N/A | High risk | Low risk |
| 18 | Non-recurrent | 0 | 55-59 | N/A | High risk | Low risk |
| 19 | Recurrent (<5 years) | 1 | 55-59 | 1553 | Low risk | Low risk |
| 20 | Non-recurrent | 0 | 60-64 | N/A | High risk | Low risk |
| 21 | Non-recurrent | 0 | 55-59 | N/A | Low risk | Low risk |
| 22 | Recurrent (<5 years) | 1 | 70-74 | 182 | Low risk | High risk |
| 23 | Non-recurrent | 0 | 70-74 | N/A | Low risk | Low risk |
| 24 | Recurrent (<5 years) | 1 | 50-54 | 214 | Low risk | High risk |
| 25 | Non-recurrent | 0 | 60-64 | N/A | High risk | Low risk |
| 26 | Non-recurrent | 0 | 65-69 | N/A | High risk | High risk |
| 27 | Non-recurrent | 0 | 55-59 | N/A | Low risk | Low risk |
| 28 | Recurrent (<5 years) | 1 | 70-74 | 1065 | High risk | High risk |
| 29 | Non-recurrent | 0 | 55-59 | N/A | High risk | High risk |
| 30 | Non-recurrent | 0 | 55-59 | N/A | Low risk | Low risk |
| 31 | Non-recurrent | 0 | 50-54 | N/A | High risk | Low risk |
| 32 | Recurrent (<5 years) | 1 | 55-59 | Data not available | Low risk | High risk |
| 33 | Recurrent (>5 years) | 0 | 55-59 | Data not available | High risk | Low risk |
| 34 | Recurrent (<5 years) | 1 | 65-69 | 457 | High risk | High risk |

| | | | | | | |
|---|---|---|---|---|---|---|
| 35 | Recurrent (<5 years) | 1 | 60-64 | Data not available | High risk | High risk |
| 36 | Non-recurrent | 0 | 55-59 | N/A | Low risk | Low risk |
| 37 | Recurrent (<5 years) | 1 | 50-54 | 1216 | High risk | High risk |
| 38 | Recurrent (<5 years) | 1 | 55-59 | 182 | High risk | High risk |
| 39 | Recurrent (<5 years) | 1 | 45-49 | 912 | High risk | High risk |
| 40 | Recurrent (<5 years) | 1 | 60-64 | Data not available | High risk | High risk |
| 41 | Recurrent (<5 years) | 1 | 65-69 | Data not available | High risk | High risk |
| 42 | Recurrent (<5 years) | 1 | 60-64 | Data not available | Low risk | High risk |
| 43 | Recurrent (<5 years) | 1 | 60-64 | Data not available | High risk | High risk |
| 44 | Recurrent (<5 years) | 1 | 60-64 | Data not available | High risk | High risk |
| 45 | Recurrent (<5 years) | 1 | 60-64 | 1096 | High risk | Low risk |
| 46 | Non-recurrent | 0 | 50-54 | N/A | Low risk | High risk |
| 47 | Recurrent (<5 years) | 1 | 70-74 | Data not available | Low risk | High risk |
| 48 | Non-recurrent | 0 | 65-69 | N/A | Low risk | Low risk |
| 49 | Non-recurrent | 0 | 50-54 | N/A | High risk | Low risk |
| 50 | Non-recurrent | 0 | 60-64 | N/A | Low risk | Low risk |

[1] 1 = cases with BCR under 5 years, 0 = all other cases.

[2] This stratification is based on a threshold of 0.5 applied to the multi-parameter classification model's output (which ranges from 0-1).

**Supplementary Table 4 | Comparison of 3D and 2.5D image translation in terms of the tissue volume that must be redundantly processed due to GPU limitations (which limit the 3D block sizes)**

| | *vox2vox* (full 3D) | *ITAS3D* (2.5D) |
|---|---|---|
| Biopsy size (pixels) | 14000 × 1024 × 712 | |
| Sub-divided block size, limited by GPU memory (pixels) | 173 × 133 × 115 (NVIDIA GeForce RTX 2080 Ti) | 1024 × 1024 × 712 (NVIDIA Tesla P100) |
| Overlap percentage | 25% | |
| Total processed volume (# of pixels) including overlap | $2.5 \times 10^{10}$ | $1.3 \times 10^{10}$ |
| Redundantly processed volume (# of pixels) | $1.4 \times 10^{10}$ | $3.2 \times 10^{9}$ |
| Percentage of biopsy volume that is redundantly processed | 142.5% | 31.7% |

**Supplementary Videos**

**Supplementary Video 1 | Comparison of synthetic-CK8 outputs generated by models trained with and without the coarse-to-fine training strategy.** A depth-stack (z-stack) video shows (from left to right): a real-CK8 dataset (ground-truth), a synthetic-CK8 dataset generated by a model trained with the coarse-to-fine strategy, and a synthetic-CK8 dataset generated by a model trained without the coarse-to-fine strategy. Two different tissue regions are shown (top and bottom rows). The results show no major qualitative or quantitative differences (3D SSIM) in image quality, but the training time is approximately doubled with the coarse-to-fine strategy. Scale bar: 100 μm.

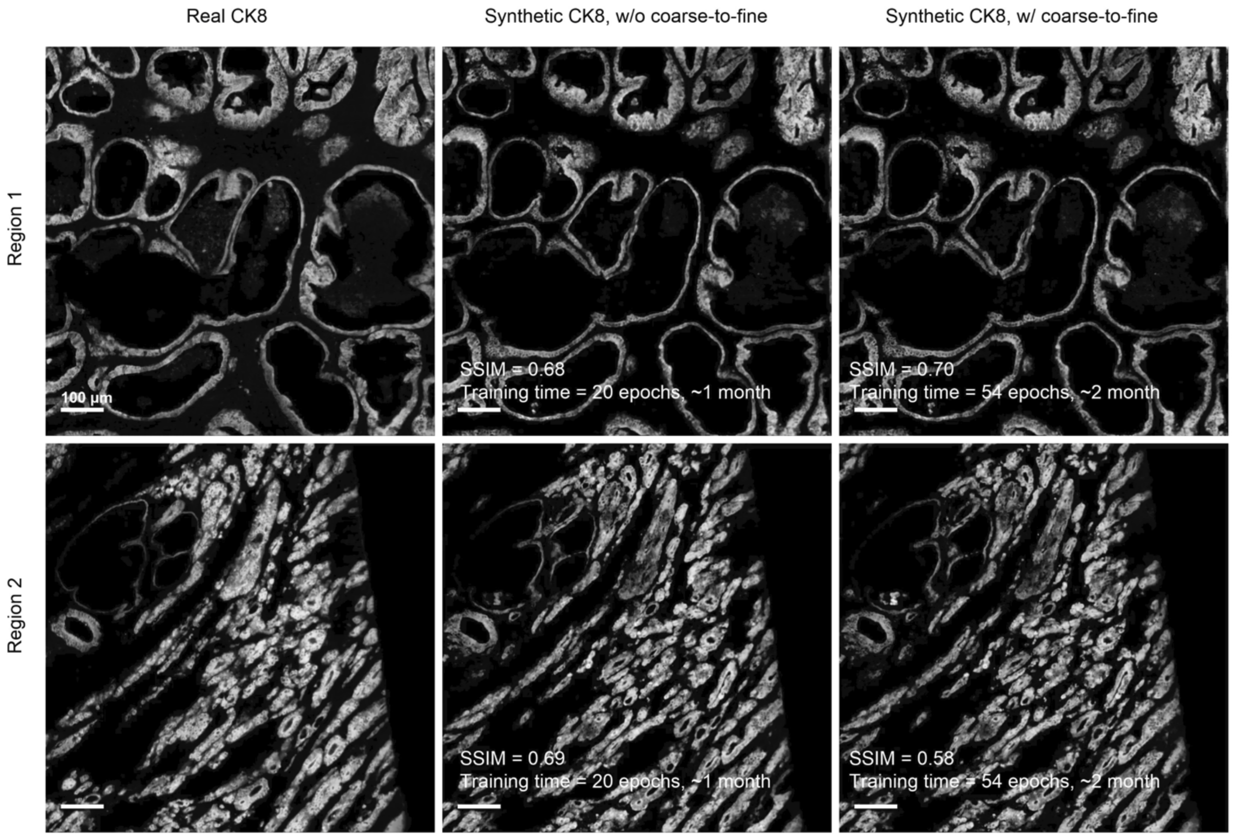


Videos are provided as separate files (a screenshot is shown above)

**Supplementary Video 2 | Depth sequence of the benign and cancerous 3D regions shown in Fig. 3.** Columns from left to right: false-colored H&E images, false-colored synthetic-CK8 images (IHC appearance), and segmentation masks for the luminal epithelium (in red), lumen (in yellow), and stroma (in gray). Scale bar: 100 μm.

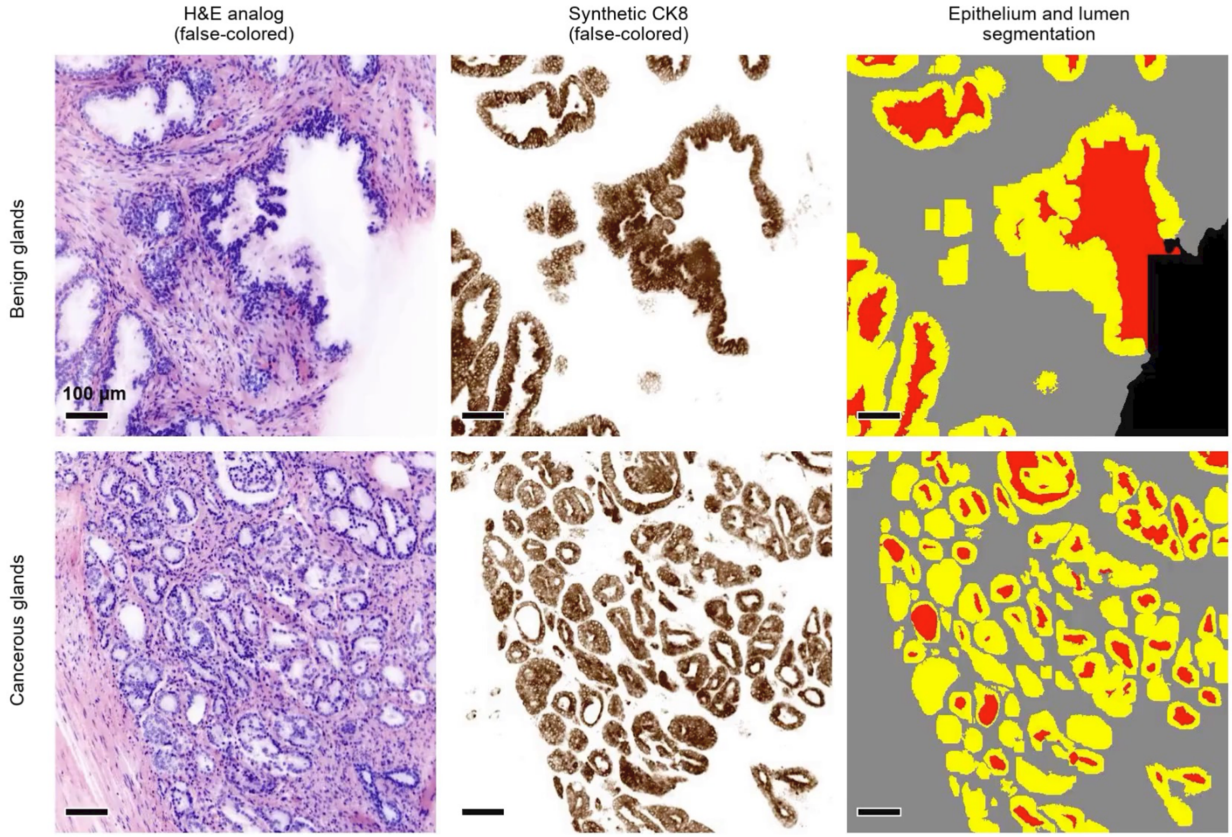


Videos are provided as separate files (a screenshot is shown above)

**Supplementary Video 3 | Depth sequence of a false-colored H&E-analog dataset, a corresponding CK8-IHC dataset, and a gland-segmentation mask for a 3D prostate biopsy.** In the segmentation mask, yellow regions represent the luminal epithelium, red regions represent the lumen, and gray regions represent the stroma. Scale bar: 100 µm.

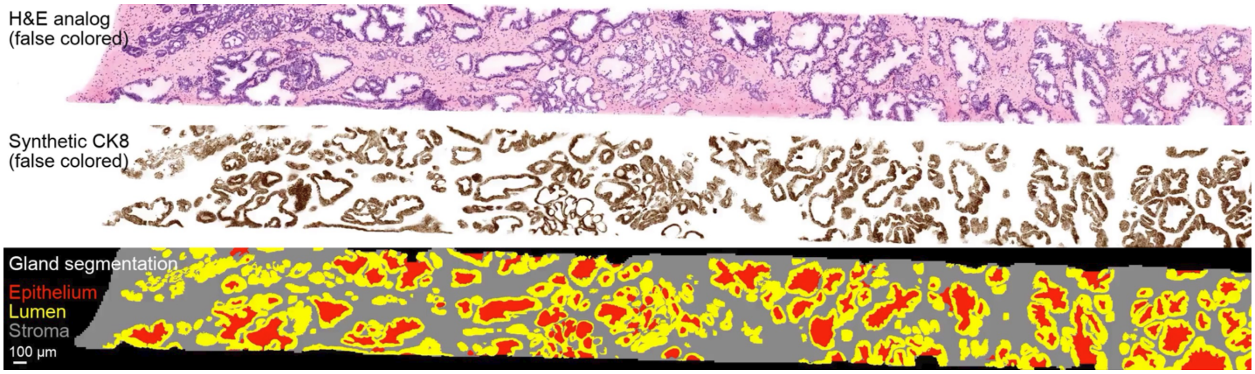


Videos are provided as separate files (a screenshot is shown above)

**Supplementary Video 4 | Comparison between 2D image translation and 2.5D image translation.** A depth-stack (z-stack) video shows (from left to right): a false-colored H&E-analog dataset (input), real-CK8 IF (ground-truth), synthetic-CK8 IF generated with 2D image translation (output), and synthetic-CK8 IF generated with 2.5D image translation (output). Two different tissue regions are shown (top and bottom rows). Compared to 2D image translation, the 2.5D image translation significantly enhances the spatial continuity between different levels as a function of depth. Scale bar: 100 µm.

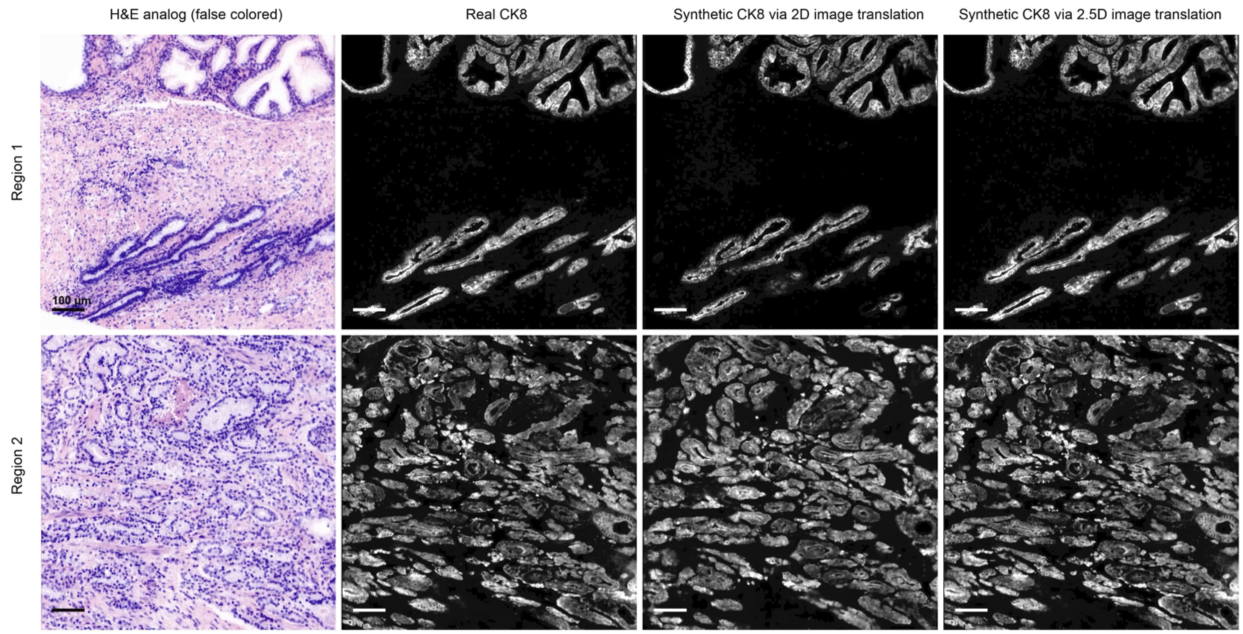


Videos are provided as separate files (a screenshot is shown above)

**Supplementary Video 5 | Comparison of segmentation results with ITAS3D and two baseline methods.** Top, front, and side views (x, y, and z-stack videos) are shown of 3D segmentations for an example tissue region achieved with ITAS3D, 2D U-net (generated level-by-level along the z direction), and 3D watershed. Regions where the segmentation mask agrees with the ground-truth annotation (segmentation+, ground-truth+) are shown in white, and regions where the segmentation disagrees with the ground-truth are shown in cyan (segmentation+, ground-truth-) and magenta (segmentation-, ground-truth+). Scale bar: 100 µm.

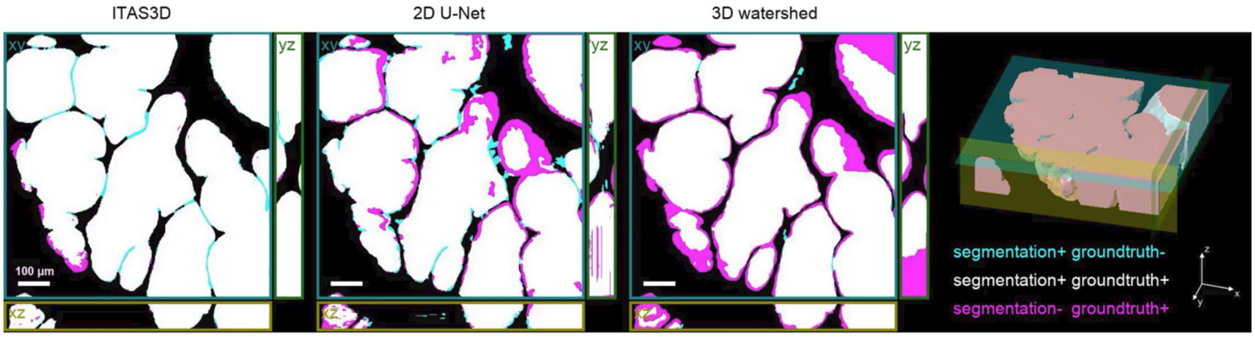


Videos are provided as separate files (a screenshot is shown above)

**Supplementary Video 6 | Volume rendering of gland segmentations and skeleton networks for benign and cancerous biopsies.**

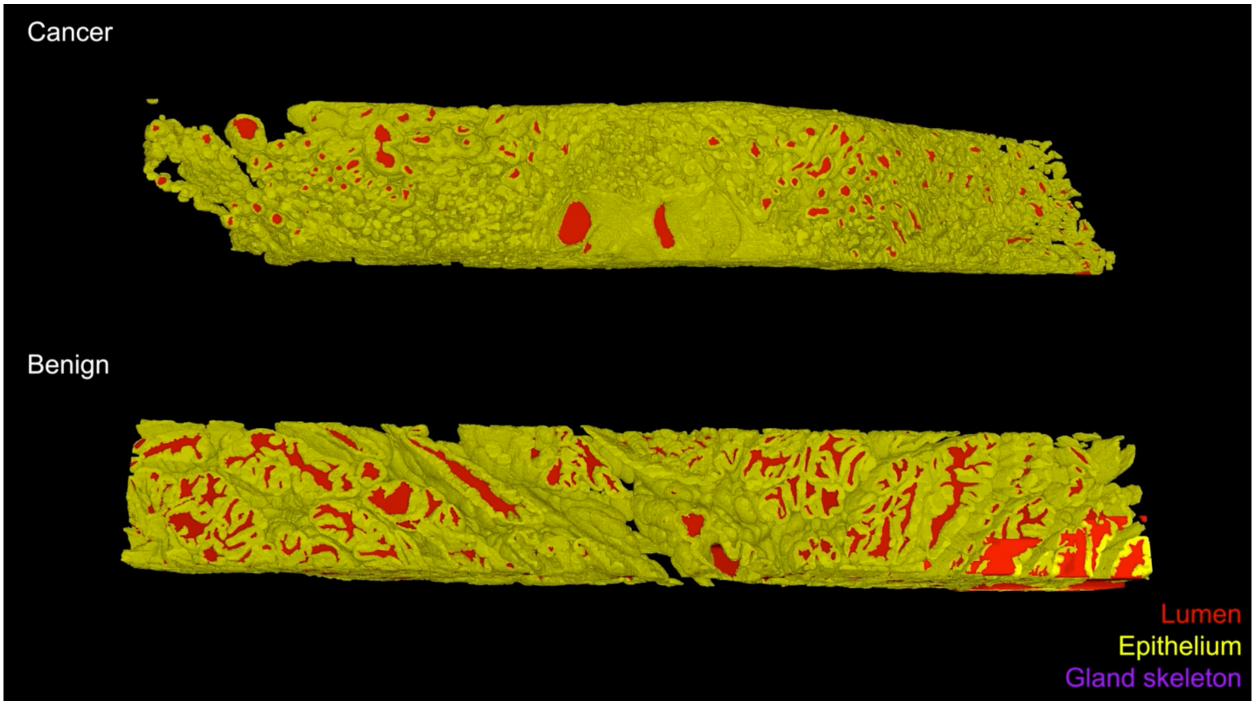


Videos are provided as separate files (a screenshot is shown above)

**Supplementary Video 7 | Video summary of ITAS3D-enabled PCa gland analysis for whole biopsies.**

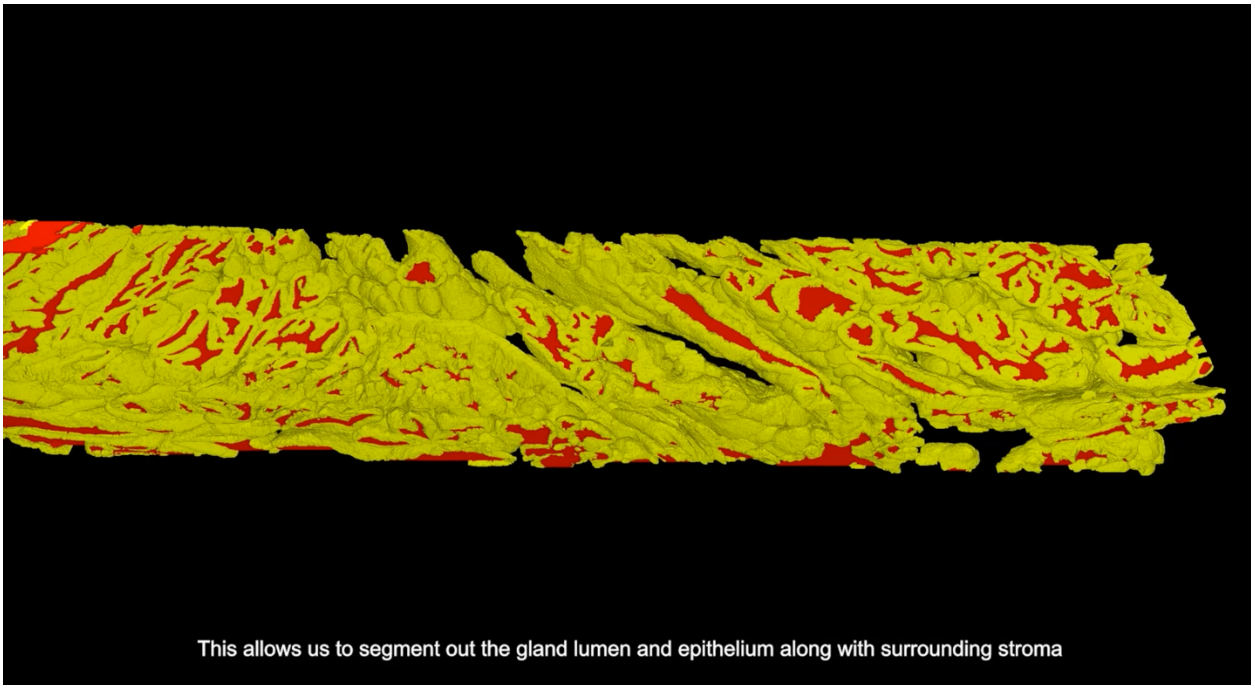


Videos are provided as separate files (a screenshot is shown above)
